# Lifestyle physical activity intensity and rapid-rate non-sustained ventricular tachycardia in arrhythmogenic cardiomyopathy

**DOI:** 10.1101/2022.07.27.22278124

**Authors:** Javier Ramos-Maqueda, Jairo H. Migueles, María Molina-Jiménez, David Ruiz-González, Eva Cabrera-Borrego, Amalio Ruiz-Salas, Alberto Soriano-Maldonado, Juan Jiménez-Jáimez

## Abstract

**Objectives:** This study aimed to investigate the association of accelerometer-measured lifestyle physical activity with the incidence of rapid-rate non-sustained ventricular tachycardias (RR-NSVT) during a 30-day recording period in AC patients.

**Methods:** This multicenter, observational study enrolled 72 AC patients, including right, left-, and biventricular forms of the disease, with underlying desmosomal and non desmosomal mutations. Lifestyle physical activity was objectively monitored with accelerometers (i.e., movement sensors), and RR-NSVT were identified from a textile Holter electrocardiogram (ECG) for 30 days as those faster than 188 bpm and longer than 18 beats.

**Results:** Sixty-three AC patients (38±17.6 years, 57% men) provided data on physical activity and ventricular tachycardias. A total of 17 patients experienced 1 ≥ RR-NSVTs, and a total of 35 events were recorded. Participants presenting RR-NSVTs during the 30-day measurement (n=17) did not perform more lifestyle physical activity (*P*=0.734), neither more activities of moderate-to-vigorous intensity (*P*=0.245) than their peers without RR-NSVTs in this period. Likewise, participants with RR-NSVTs did not increase their activity level, neither the activities of any intensity (−5 min/day on average) nor those of moderate-to-vigorous intensity (+2 min/day on average) in the days of RR-NSVTs occurrence. Finally, we observed that around half of the RR-NSVTs recorded in the 30 days occurred at rest (i.e., 19 RR-NSVTs), and the other half occurred after or during physical activity (i.e.,16 RR-NSVTs).

**Conclusion:** These findings suggest that lifestyle physical activity, mainly of light and moderate intensity, does not increase the risk for RR-NSVTs in AC patients.

**Summary box:** *What is already known:*

- Arrhythmogenic cardiomyopathy (AC) is associated to sport-triggered sudden cardiac death
- Competitive sports and high-intensity exercise are contraindicated in AC patients
- Rapid-rate non-sustained ventricular arrhythmias (RR-NSVT) are a surrogate of sudden cardiac death in this population

*What this study adds:*

- Conscious investigation on the relationship between lifestyle physical activity of different intensities with ventricular arrhythmias in AC patients
- Accelerometer-measured lifestyle physical activity, mainly of light and moderate intensity, seem not to be associated with the occurrence of RR-NSVTs

*How this study might affect clinical practice in the future:*

- This study provides strong evidence for the promotion of lifestyle physical activity among AC patients to ensure they obtain the multiple benefits and risk reductions associated with physical activity

## Introduction

Arrhythmogenic cardiomyopathy (AC) is a genetic cardiac condition characterized by fibrofatty replacement of the ventricular myocardium in one or both ventricles [1]. This provides the anatomical substrate for ventricular tachycardias and ventricular fibrillation, as well as increased risk for sudden cardiac death [2]. The underlying genetic etiology is expanding with the identification of non-desmosomal targets such as intermediate filaments, ion channels and nuclear envelope proteins, although evidence on these variants is still emerging [1,3]. The presence of rapid-rate non-sustained ventricular tachycardia (RR-NSVT) predicts appropriate future implantable cardioverter-defibrillator (ICD) shocks, interpreted as a surrogate marker for sudden cardiac death [4]. Thus, RR-NSVT has been classified as an important risk factor for sudden cardiac death in AC patients [5].

Competitive sports are contraindicated in AC patients since they increase the risk of SCD [6–8]. Less evidence is available on the role of lifestyle physical activities. Lifestyle physical activities may reach light (i.e., 1.5-2.9 times the resting energy expenditure or metabolic equivalents [METs]), moderate (i.e., ≥3 METs) or even vigorous (i.e., ≥6 METs) intensities [9]. AC patients are exposed to these lifestyle activities in their daily life (e.g., when commuting, working, or in their leisure time) [10], and there are not evidence-based guidelines about whether lifestyle activities of any intensity should be discouraged. Activities of light-to-moderate intensity are recommended in this population based on previous observational studies [6–8,11]. However, these studies are limited by their retrospective designs and the use of self-reported exercise and sport participation measures, which are of questionable validity and reliability [12–14].

Modern accelerometers (i.e., movement sensors) allow for long-term and objective monitoring of physical activity, thus allowing to assess the risk associated with lifestyle physical activity in AC patients, as well as the observation of the amount of movement the participant was doing at the exact moment in which a RR-NSVT occurred. Therefore, this study aimed to evaluate the association of device-measured lifestyle physical activity with the incidence of RR-NSVT in AC patients including a wide spectrum of the disease with high prevalence of left and biventricular forms. Our hypotheses were: (i) participants experiencing RR-NSVTs during the physical activity recording perform more physical activity than participants not experiencing RR-NSVTs (person-level analysis); (ii) those participants experiencing RR-NSVTs during the recording change their lifestyle physical activity in the days in which they have a RR-NSVT (day-level analysis); and, (iii) RR-NSVTs mostly occur during or after the performance of physical activity (epoch-level analysis).

## Methods

### Study design and population

AC probands and family members (including silent carriers) were enrolled from two Spanish centers (Virgen de las Nieves and Virgen de la Victoria University Hospitals Referral Inherited Heart Diseases Clinics) in this multicenter, observational study. The baseline assessments and a continuous recording of physical activity and ventricular arrythmias for 30 days were conducted from May 2019 to March 2021. The study population comprised 72 AC patients diagnosed based on the 2010 international diagnostic Padua criteria, including either arrhythmogenic right, left ventricular cardiomyopathy, or biventricular forms [15]. Patients aged less than 14 years, and those who could not wear the wearable devices (i.e., accelerometers and textile electrocardiogram [ECG]) because of skin allergies or job incompatibilities were excluded. Of the 72 AC patients initially recruited, patients wearing the accelerometers and the textile ECG for less than 14 days were excluded (n=9), resulting in an analytical sample of 63 patients. All patients signed an informed consent, and the study was approved by the Local Ethics Committee.

### Procedures

All patients were clinically and genetically characterized with ECG, advanced cardiac imaging, and next generation sequencing AC panels at baseline. Left and right ventricular volume and function was analyzed according to standard guidelines and AC was classified in left, right or biventricular form. Those measurements were followed by a 30-day monitoring of the physical activity with movement sensors (Axivity AX3, OmGui, Newcastle University) and ventricular arrythmias with a textile ECG Holter (Holter Nuubo ECG PLATFORM^®^). Participants were asked to maintain their usual daily activities during the recording time, and to wear the devices as much as possible (i.e., except for showers and water-based activities). The movement sensors and ECG Holter data were then processed by staff members masked the other device information (i.e., staff in charge of the accelerometer data processing did not have access to the Holter information and vice versa).

### Measurements

#### Physical activity

The lifestyle physical activity was continuously monitored with a movement sensor worn on the non-dominant wrist (Axivity AX3, OmGui, Newcastle University) for 30 consecutive days. Participants wore the sensors during the whole day and night and were instructed to only remove the devices for water-based activities (e.g., swimming). The accelerometers were set to record accelerations at 25 Hz to ensure the battery life and storage capacity during the whole period. Raw acceleration values were then downloaded in the OmGui open-source software (OmGui, Newcastle University) and saved in cwa format. These files were then processed in the open-source software GGIR [16]. The data processing pipeline included: (i) auto-calibration of the data according to the local gravity [17]; (ii) cleaning and aggregation of the acceleration values over 5-second epochs [18]; (iii) detection and imputation of the non-wear time based on an automated algorithm [19]; (iv) classification of waking and sleeping times based on the variability of the arm position [20,21]; (v) classification of the detected awake time as sedentary, light, moderate, and vigorous physical activity intensity based on previously calibrated acceleration thresholds [22,23].

Activities of vigorous intensity were rarely performed by the AC patients included in this study (i.e., 80% of participants performed less than 1 min/day in vigorous intensity activities, and 5.7 min/day was the maximum value recorded) [10]. Thus, we merged the time in activities of moderate and vigorous intensity into one category (i.e., moderate-to-vigorous physical activity) for person- and day-level analyses. Additionally, we also combined all activities of any intensity (i.e., including light, moderate, and vigorous intensity) for analyses. All days in which the participants wore the devices for at least 16 hours were included in the analyses.

#### Ventricular tachycardia

Ventricular tachycardias were recorded during a 30-day period with the Holter Nuubo ECG PLATFORM^®^. Sustained ventricular tachycardias were defined as those of at least 100 bpm for more than 30 seconds, while RR-NSVTs were defined as at least 100 bpm and less than 30 seconds. Episodes not fulfilling both criteria were not used for the analysis. The RR-NSVTs have been previously associated with an increased risk of appropriate ICD shocks as a surrogate of sudden cardiac death [24]. An independent electrophysiologist blinded for the patients’ medical history and their exercise level reviewed the ECG Holter and recorded RR-NSVT.

### Statistical analysis

Participants’ descriptive characteristics at baseline were summarized as mean and standard deviation (SD), or frequencies and percentages, as appropriate. First, in the person-level analysis, we compared the average daily time the participants engaged in activities of moderate-to-vigorous intensity, as well as activities of any intensity in those patients who experienced at least one RR-NSVT versus their peers who did not-record any RR-NSVT during the 30-day recording. For such purpose, we performed analysis of covariance (ANCOVA) models adjusted for age, sex, implanted ICD, and left-ventricular ejection fraction. Second, the day-level analysis evaluated whether the AC patients experiencing RR-NSVTs during the 30-day recording period performed more physical activity in those days in which the RR-NSVTs occurred (compared with the days without RR-NSVT occurrence). For such purpose, we run a linear mixed model to compare the lifestyle physical activity indicator (i.e., moderate-to-vigorous intensity or any intensity activities) in the days the participants had an event versus the days the participants did not (fixed factor) and considering the patients IDs as random factor. Random and fixed intercepts were defined to investigate the within-group (occurrence or not of RR-NSVT) and within-patient effect of physical activity. Finally, at epoch-level, two independent raters visually classified whether the RR-NSVTs occurred during or after the performance of physical activity, or at rest. The two raters agreed in 97% of the classifications, and the agreement reach 100% after a consensus meeting. All the analyses were performed in R version 4.2.0 and the statistical significance level was set at *P*<0.05.

## Results

### Baseline characteristics of patients

**Table 1** presents the baseline characteristics of the patients included in the analyses (n=63). Most of the patients presented with a left or biventricular form of AC due to a non-desmosomal gene mutation, with a mean LVEF of 50.39 ± 11.92 %. There was a high prevalence of familial SCD and prior ventricular arrhythmias, so more than half of the patients carried an ICD for primary and secondary prevention of SCD. The most prevalent underlying mutated genes were DES, DSP and FLNC. The patients performed an average of 290 (SD=123) min/day of physical activity in their daily life, being 22 (SD=20) min/day activities of moderate-to-vigorous intensity. A total of 35 RR-NSVTs were recorded during the 30-day measurements, with 17 participants presenting at least one RR-NSVT during the data monitoring. However, no sustained ventricular tachycardias were observed during the recording period.

**Table 1.** Characteristics of patients (n=63).

| Characteristic | Value |
| --- | --- |
| *Sociodemographic* |  |
| Male sex, n (%) | 36 (57) |
| Age (y) | 48.0 (17.6) |
| *Genetics* |  |
| Desmosomal mutations, n (%) | 23 (36.5) |
| DSP | 10 (15.9) |
| DSG2 | 5 (7.9) |
| PKP2 | 8 (12.7) |
| Non desmosomomal mutations, n (%) | 29 (46) |
| FLNC | 14 (22.2) |
| DES | 8 (12.7) |
| TMEM43 | 5 (7.9) |
| LMNA | 2 (3.2) |
| Gene elusive, n (%) | 11 (17.5) |
| *Phenotypic form* |  |
| ARVC, n (%) | 24 (38.1) |
| ALVC, n (%) | 32 (50.8) |
| Biventricular AC, n (%) | 7 (11.1) |
| *Clinical features* |  |
| Left ventricular ejection fraction, n (%) |  |
| > 52% | 34 (54) |
| ≤ 52% | 29 (46) |
| VT/VF history | 21 (33.3) |
| ICD, n (%) | 36 (57) |
| Appropriate ICD shocks history | 11 (17.5) |
| *Physical activity* |  |
| Moderate-to-vigorous intensity (min/day) | 20.9 (16.3) |
| Total physical activity (min/day) | 290.2 (123.2) |
| *Ventricular Tachycardias, n* |  |
| Sustained | 0 |
| RR-NSVT | 35 |

Mean (SD) unless otherwise stated.
AC, arrhythmogenic cardiomyopathy; ALVC, arrhythmogenic left ventricular cardiomyopathy; ARVC, arrhythmogenic right ventricular cardiomyopathy; DES, desmin; DSG2, desmoglein-2; DSP, desmoplakin; FLNC, filamin C; ICD, implantable cardioverter defibrillator; LMNA, Lamin A/C; LVEF, left ventricular ejection fraction; RR-NSVT, rapid-rate non-sustained ventricular tachycardia; PKP2, plakophilin-2; TMEM43, transmembrane protein 43; VT/VF, ventricular tachycardia/ventricular fibrillation.

### Person-level associations

At person level, those AC patients who experienced at least one RR-NSVT in the 30-day measurement had not significantly different levels of physical activity than their peers who did not experience any RR-NSVT (**Figure 1**). Specifically, those with RR-NSVTs performed 6 minutes per day less of activities of moderate-to-vigorous intensity (17 [CI^95%^: 9 to 25] versus 23 [CI^95%^: 18 to 28] min/day, *P*=0.245) and 13 minutes per day less of total physical activity (283 [CI^95%^: 220 to 346] versus 296 [CI^95%^: 259 to 334] min/day, *P*=0.720) after adjusting for age, sex, implanted ICD, and left-ventricular ejection fraction.

**Figure 1.**
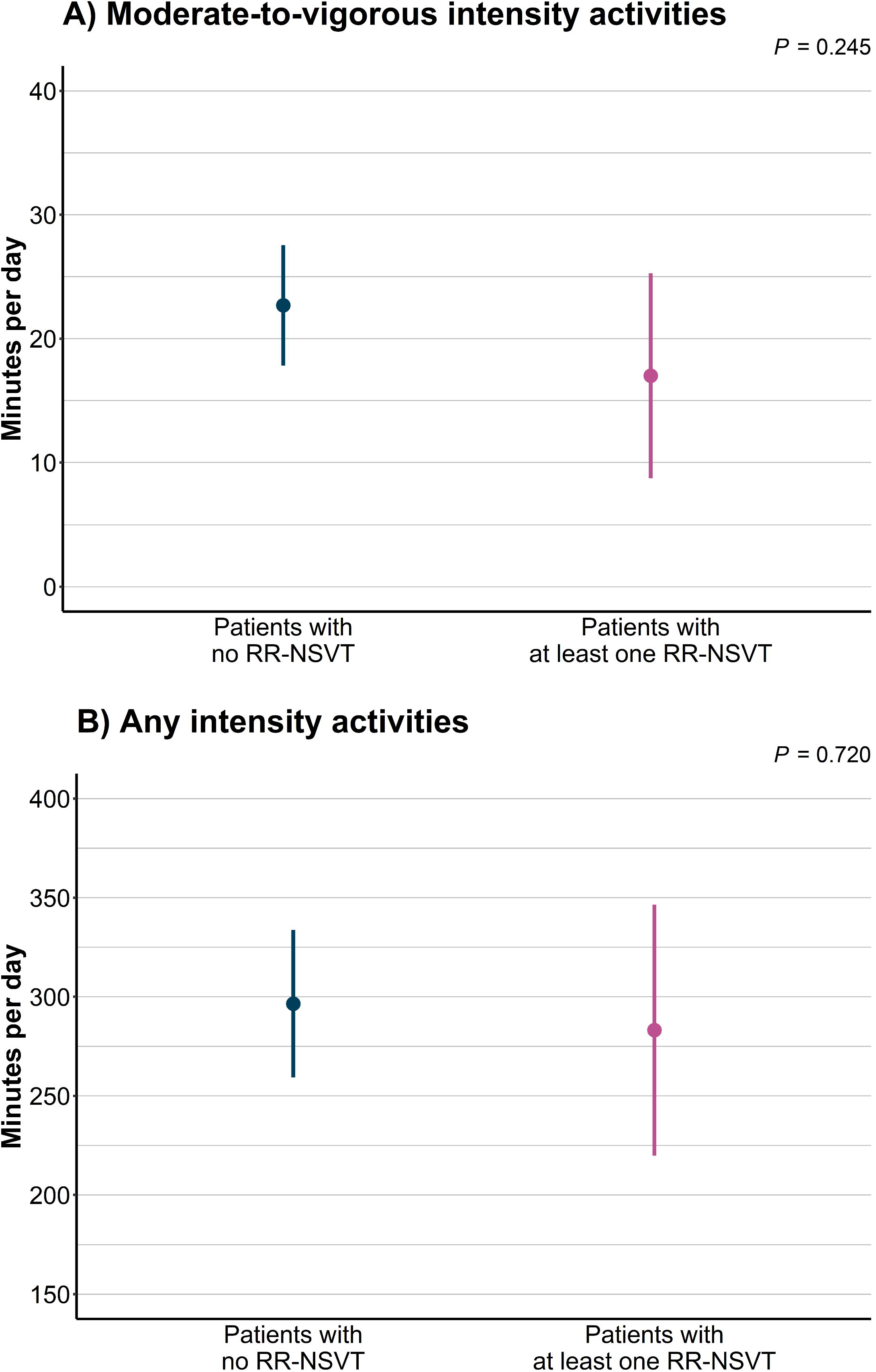
Estimated marginal means and 95% confidence intervals for the comparison of the daily time in (A) moderate-to-vigorous intensity activities and (B) any intensity activities between participants having at least one RR-NSVT during the 30-day recording period (n=17) and their peers who did not have any (n=46). Note: *P* value from ANCOVA model adjusted for age, sex, implanted ICD, and left ventricular ejection fraction. RR-NSVT: rapid-rate non-sustained ventricular tachycardia, ICD: implantable cardioverter-defibrillator.

### Day-level associations

At day level, those AC patients with RR-NSVTs during the recording (n=17) did not show different levels of physical activity in the days when the RR-NSVTs occurred compared with their days without any RR-NSVT (**Figure 2**). These patients performed 2 minutes per day more of moderate-to-vigorous intensity activities (17 [CI^95%^: 9 to 25] versus 15 [CI^95%^: 9 to 20] min/day, *P*=0.591) and 5 minutes less of total physical activity (266 [CI^95%^: 237 to 296] versus 271 [CI^95%^: 223 to 320] min/day, *P*=0.757) in the days with RR-NSVTs. At individual level, most participants showed similar patterns of moderate-to-vigorous physical activity (±6 min/day), except for 3 patients that increased the time in activities of moderate-to-vigorous intensity (+9, +12, and +16 min/day). All patients showed similar levels of total physical activity in the days they had RR-NSVTs compared to the days they did not (±20 min/day), except for one patient that spent 44 minutes less per day in the days with RR-NSVTs.

**Figure 2.**
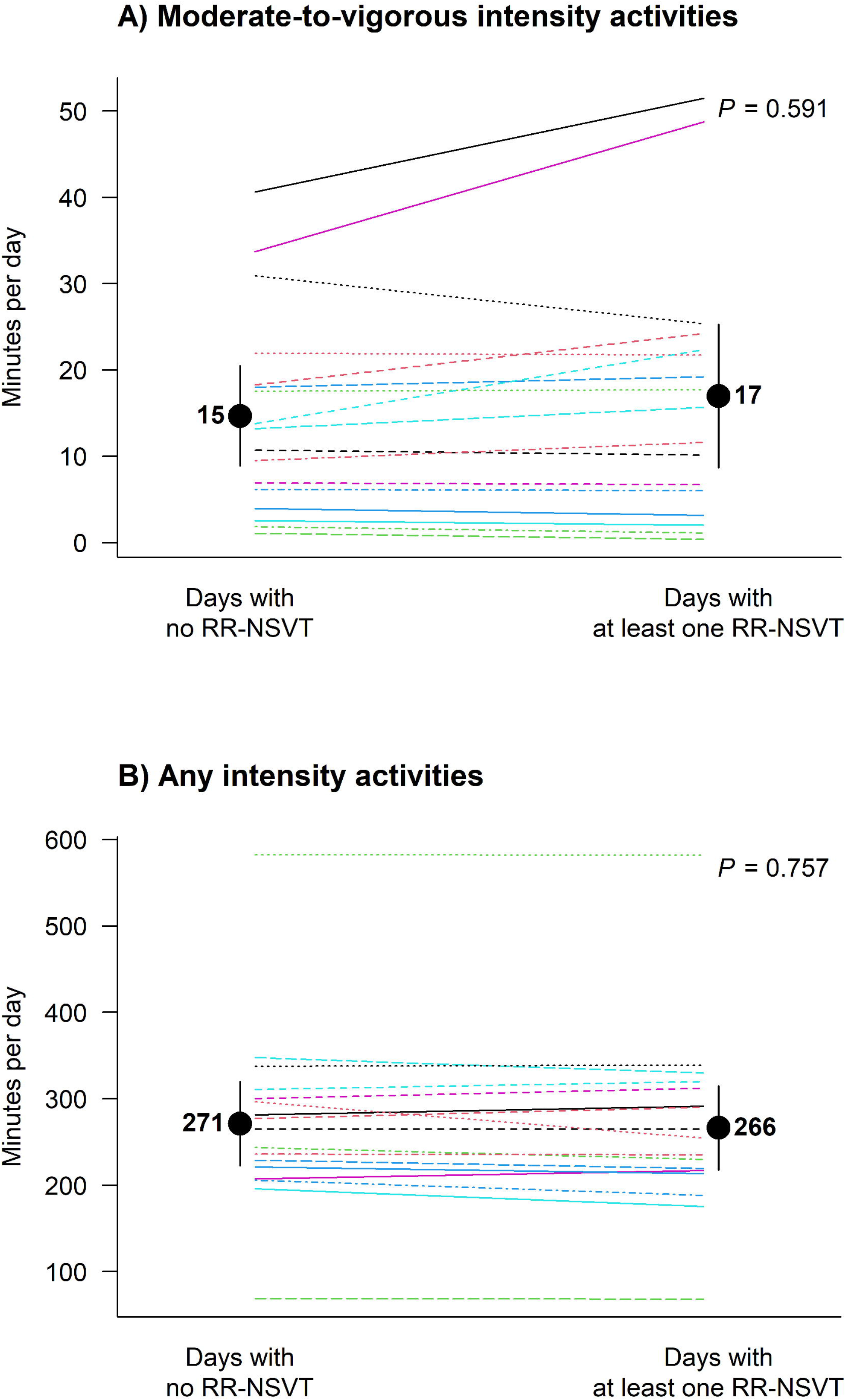
Mean differences of the daily time in (A) moderate-to-vigorous intensity activities and (B) any intensity activities between participants between the days with at least one RR-NSVT (i.e., 443 days) and the days without any (i.e., 30 days) during the 30-day recording period in those patients having at least one RR-NSVT (n=17). Note: individual and group differences estimated from mixed models with the days in which the RR-NSVT occurred (yes or no) as fixed factor and the patient IDs as random factor. RR-NSVT: rapid-rate non-sustained ventricular tachycardia.

### Epoch-level associations

Finally, at epoch level, an example of the visualizations we made and how they were classified is shown in **Figure 3** (the rest of the visualizations are presented in the **Supplement 1**). We observed that around half of the RR-NSVTs occurred at rest (i.e., 19 RR-NSVTs) during the 30-day recording, while the other half (i.e., 16 RR-NSVTs) occurred either after or during the performance of physical activity of different intensities. None of the RR-NSVTs occurred after or during the realization of vigorous physical activity.

**Figure 3.**
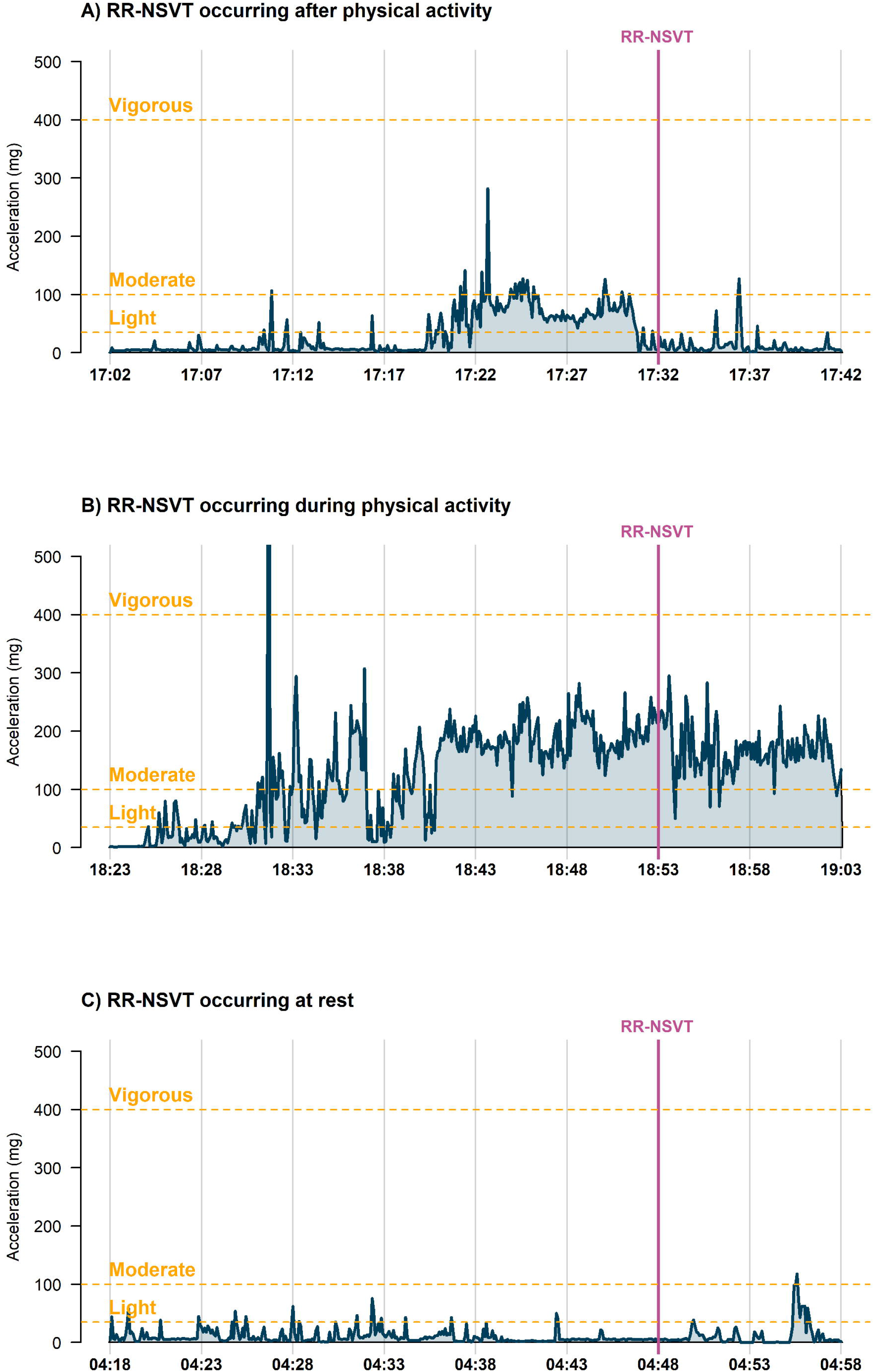
Examples of RR-NSVTs occurring (A) after performance of physical activity, (B) during performance of physical activity, and (C) at rest during the 30-day recording period. RR-NSVT: rapid-rate non-sustained ventricular tachycardia.

## Discussion

The benefits of physical activity for cardiovascular, metabolic, and mental health are widely investigated both in the general population and in people living with chronic conditions [25]. However, AC is associated to sport-triggered sudden cardiac death and, therefore, competitive sports and high-intensity exercise are contraindicated in this population [6–8,26]. Besides regular exercise and competitive sports, whether the lifestyle physical activity naturally performed by AC patients may lead to ventricular arrhythmias and, subsequently, increase the risk for sudden cardiac death remains controversial. Against our hypotheses, the main findings of this study suggest that lifestyle physical is not associated with the occurrence of RR-NSVTs in AC patients during a 30-day measurement period. Interestingly, our data suggest that lifestyle physical activity, mainly of light and moderate intensity, do not increase the arrhythmic risk in AC patients; and that these activities can be encouraged in these patients to obtain the health benefits of physical activity, which are beyond cardiovascular health [25,27].

In 2003, Corrado et al. demonstrated that 18-35-year-old athletes living with AC were at an increased risk of sudden cardiac death compared to age-matched non-athletes with AC [28]. Several subsequent studies observed that self-reported regular exercise and athletic activity are associated with more electrical or structural progression of AC [29–33], and it seems that exercise intensity has a greater influence in the development of the AC phenotype and in the arrhythmic burden than exercise duration [34]. Furthermore, Ruwald et al. found that those AC patients who reported participating in competitive/professional sports before and after diagnosis were at increased risk of ventricular tachycardia and death than those reporting either participating in recreational sports or being inactive [11]. They did not observe an increment in the risk in those patients participating in recreational sports compared to the inactive ones [11]. These studies have common limitations, as the under-representation of non-desmosomal AC patients, precluding the extrapolation of these findings to the recently defined non-desmosomal forms of AC; and the use of self-reported exercise and sports history, which is subject to recall bias, social desirability bias, and inaccuracy [13,14]. Additionally, the retrospective design of these studies might increase the recall bias in the measurement of the exercise and sports participation in the past.

Regular exercise and sports are not the only source of physical activity; AC patients may also perform physical activities of different intensities in their daily life (e.g., for transportation, work-related, or in their leisure time). The association of the lifestyle physical activity with the risk of ventricular tachycardia has not been investigated to date. In this study, we observed that lifestyle physical activity was not associated with the occurrence of RR-NSVTs in AC patients, including both desmosomal and non-desmosomal forms [10]. Specifically, we observed that: (i) AC patients with RR-NSVTs did not show higher lifestyle physical activity levels than AC patients without RR-NSVTs during our 30-day recording; (ii) AC patients with RR-NSVTs showed similar physical activity patterns in the days when the RR-NSVTs occurred compared to the days without RR-NSVT; and (iii) the RR-NSVTs did not systematically occurred after or during physical activity. It is noteworthy that AC patients did barely engage in activities of vigorous intensity, with an average of 3 min/day in this study sample and with 80% of the patients not reaching 1 min/day. Therefore, these findings mainly apply to activities of light and moderate intensity, which seem to be safe for AC patients, and it is in line with current guidelines for this population [8]. Future studies should investigate the risk of performing physical activities of higher intensities in the daily life by including AC patients with higher activity levels.

This study contributes to the existing literature by: (i) investigating the lifestyle physical activity in AC patients in relation to the arrhythmic risk, whereas previous literature had only focused on competitive sports and regular exercise; (ii) including AC patients presenting both desmosomal and non-desmosomal variants instead of only including desmosomal forms of AC as previous studies; (iii) using objective tools to quantify the physical activity, while previous research relied on self-reported retrospective methods (e.g., questionnaires, interviews) of questionable validity and reliability; (iv) the measurement of physical activity of all intensities, including light, moderate, and vigorous, instead of only relying on high-intensity regular exercise and sports participation; (v) the objective assessment of RR-NSVTs, which are considered a surrogate outcome of sudden cardiac death.

### Limitations

This study presents a number of limitations. First, the outcome was RR-NSVTs of at least 18 beats instead of sustained ventricular tachycardias or ventricular fibrillation. This decision was made under the unfeasibility of conducting longer-term continuous recordings (which would be required to observe sustained ventricular tachycardias) with both movement sensors and the textile Holter ECG. In this sense, RR-NSVTs are a well-recognized, powerful, and independent risk factor for sudden cardiac death in AC patients [5,35,36]. A longer term follow-up of the patients included in this study is planned and will provide novel information on the relationship of objectively measured lifestyle physical activity and the risk of sustained ventricular tachycardia and ventricular fibrillation in AC patients. Second, the study participants rarely engaged in vigorous intensity activities; thus, our results cannot be extrapolated to lifestyle activities of vigorous intensities. Third, the short-term follow up of the participants (30 days) might limit the clinical implications of the study; yet the objective and high-quality measurements conducted shed light on the safety of light and moderate intensity activities performed in the daily life by people living with AC.

## Conclusions

The findings of this study suggest that device-measured lifestyle physical activity, mainly of light and moderate intensity, does not increase the risk for RR-NSVTs in AC patients. This provides strong evidence to the active promotion of lifestyle physical activity among patients with AC to ensure they obtain the multiple benefits and risk reductions associated with physical activity. This study provides clinically relevant evidence for cardiologist to use in clinical practice, although future prospective studies with larger sample sizes and prospective follow up are needed to characterize the effects of lifestyle physical activity of moderate-to-vigorous intensity in AC phenotype and arrhythmic outcomes.

## Data Availability

All data produced in the present study are available upon reasonable request to the authors

## Acknowledgements

This work was funded by the Consejería de Salud y Familias, Junta de Andalucía (Ref. PIER-0231-2019). Jairo H. Migueles is supported by the Swedish Research Council for Health, Working Life and Welfare (2021-00036). This article is part of a Doctoral Thesis to be submitted by JR-M to the Clinical Medicine and Public Health Doctoral Program at the University of Granada (Spain).

