## Supplement 1 for "Lifestyle physical activity intensity and rapid-rate non-sustained ventricular tachycardia in arrhythmogenic cardiomyopathy"

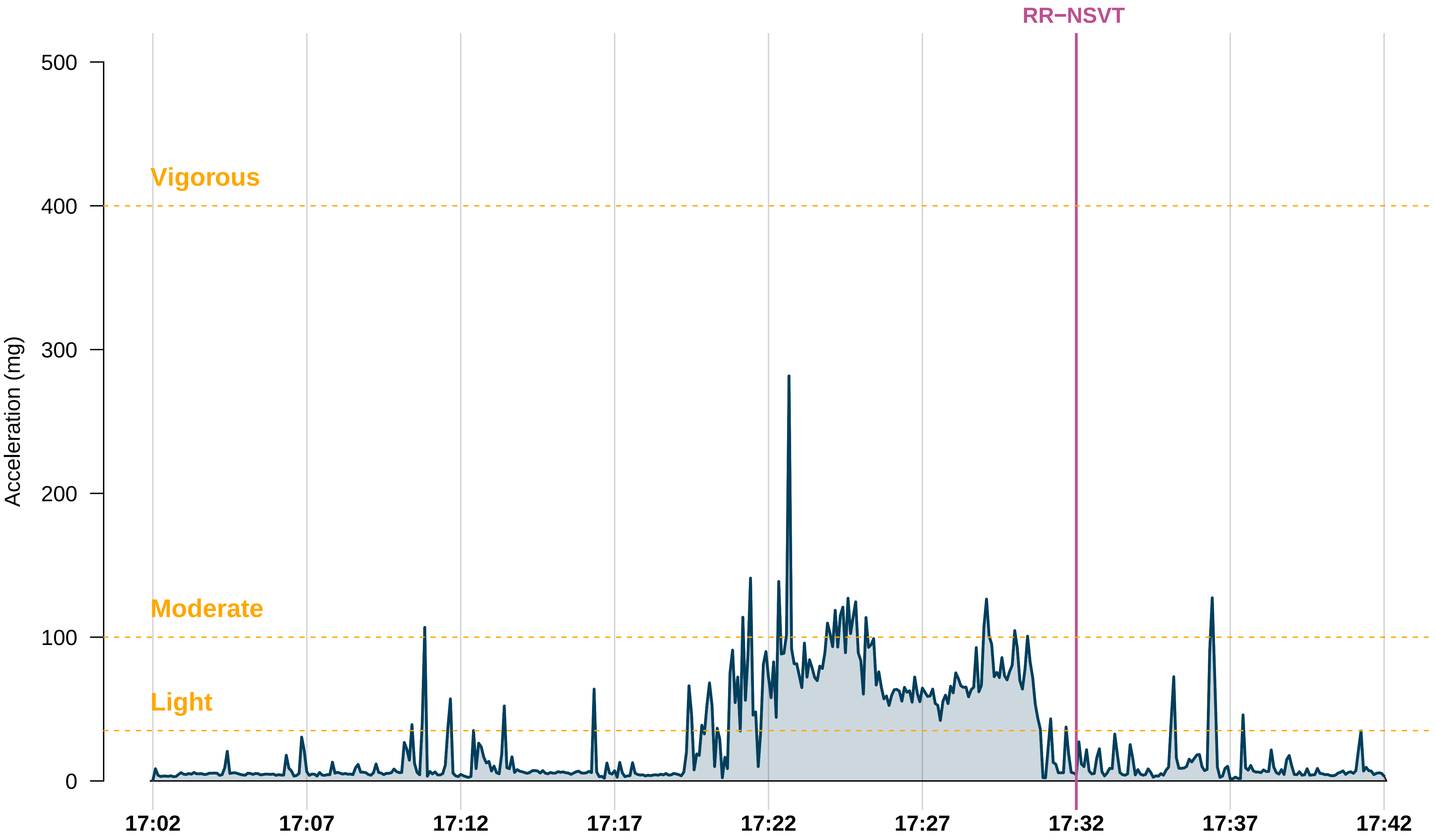
RR−NSVT
Vigorous
Moderate
Light
Acceleration (mg)
500
400
300
200
100
0
17:02
17:07
17:12
17:17
17:22
17:27
17:32
17:37
17:42

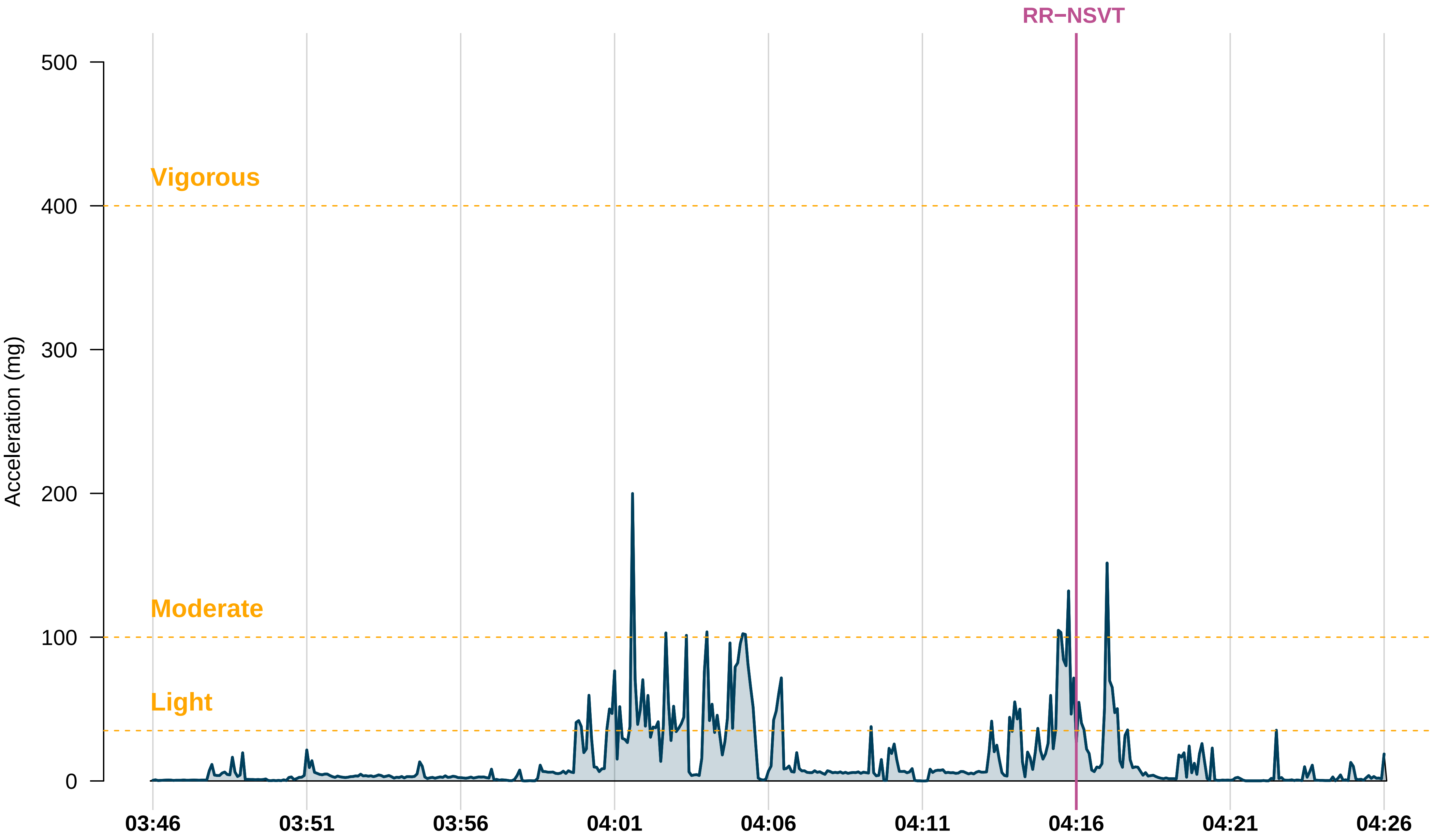

RR−NSVT
Vigorous
Moderate
Light
Acceleration (mg)
500
400
300
200
100
0
03:46
03:51
03:56
04:01
04:06
04:11
04:16
04:21
04:26

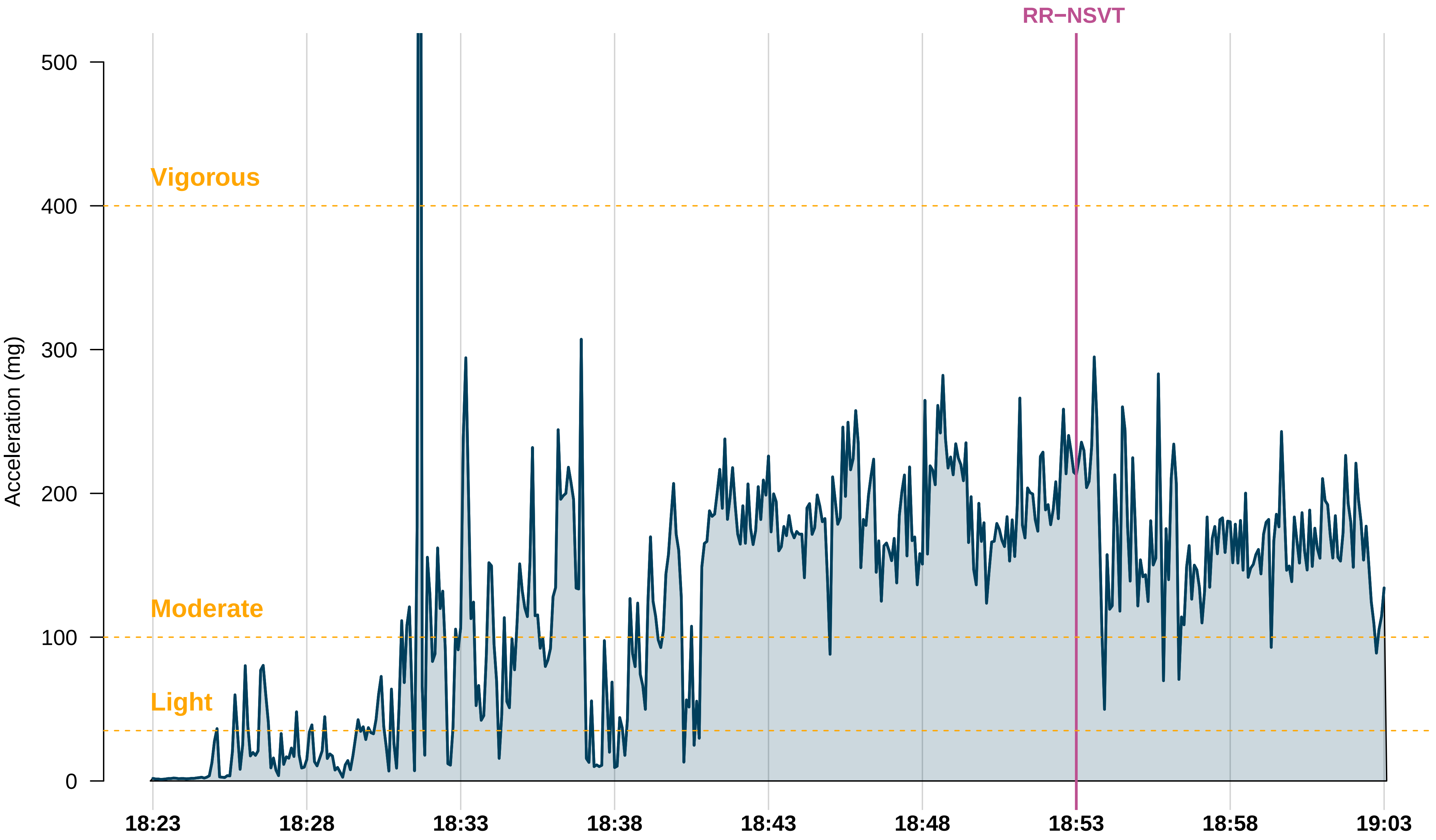
RR−NSVT
Vigorous
Moderate
Light
Acceleration (mg)
500
400
300
200
100
0
18:23
18:28
18:33
18:38
18:43
18:48
18:53
18:58
19:03

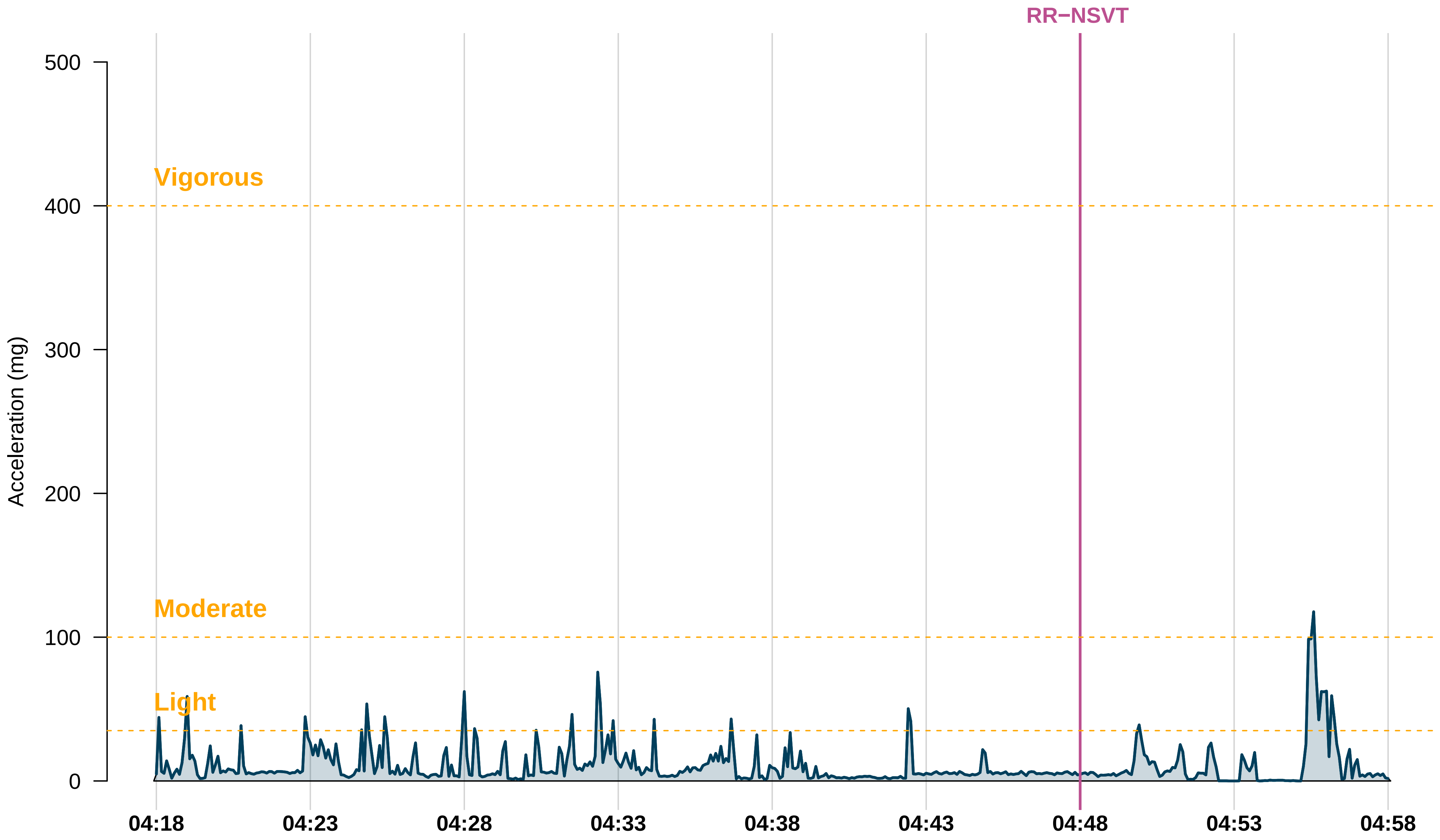

RR−NSVT
Vigorous
Moderate
Light
Acceleration (mg)
0
100
200
300
400
500
04:18
04:23
04:28
04:33
04:38
04:43
04:48
04:53
04:58

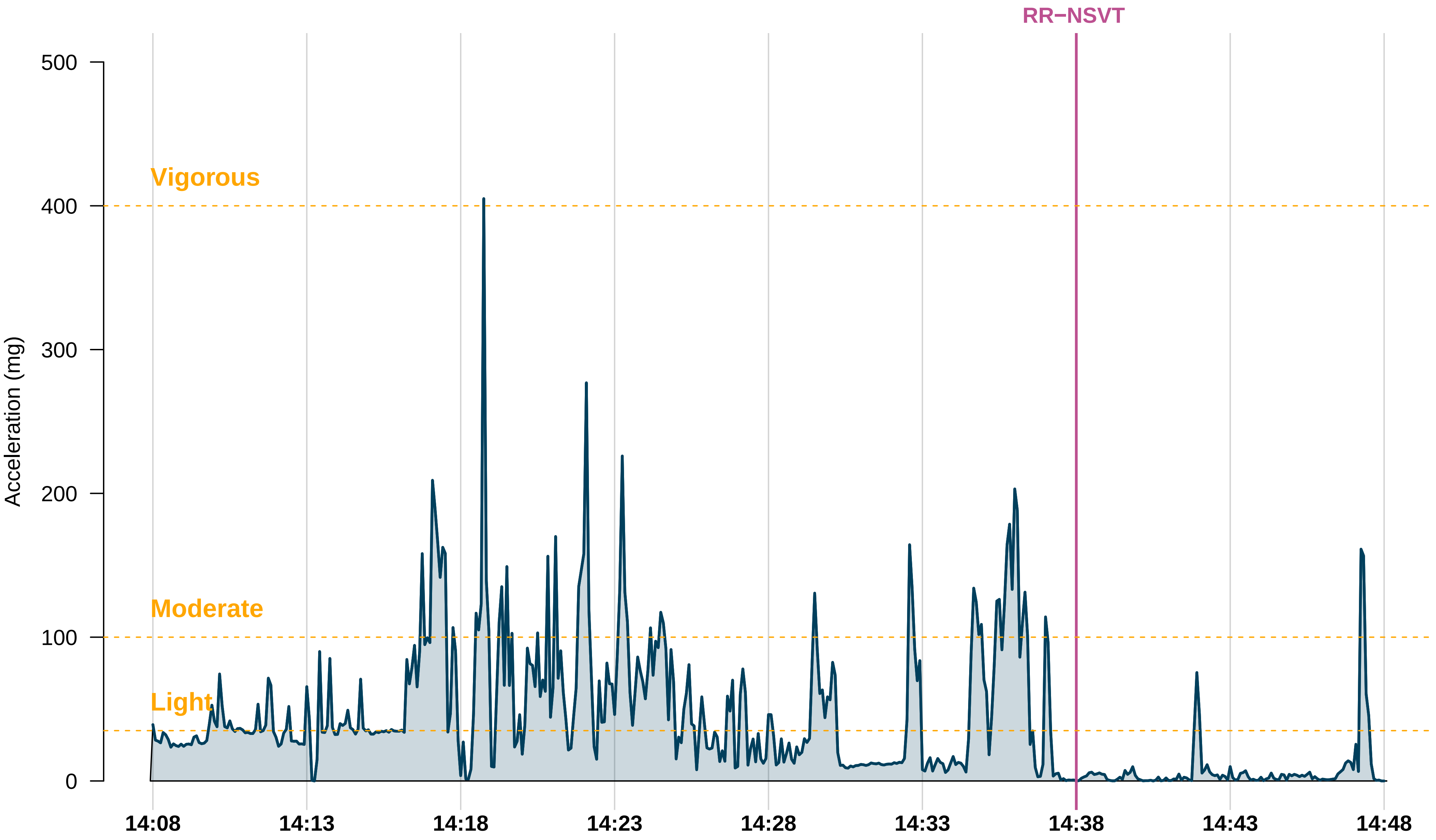

RR−NSVT
Vigorous
Moderate
Light
Acceleration (mg)
500
400
300
200
100
0
14:08
14:13
14:18
14:23
14:28
14:33
14:38
14:43
14:48

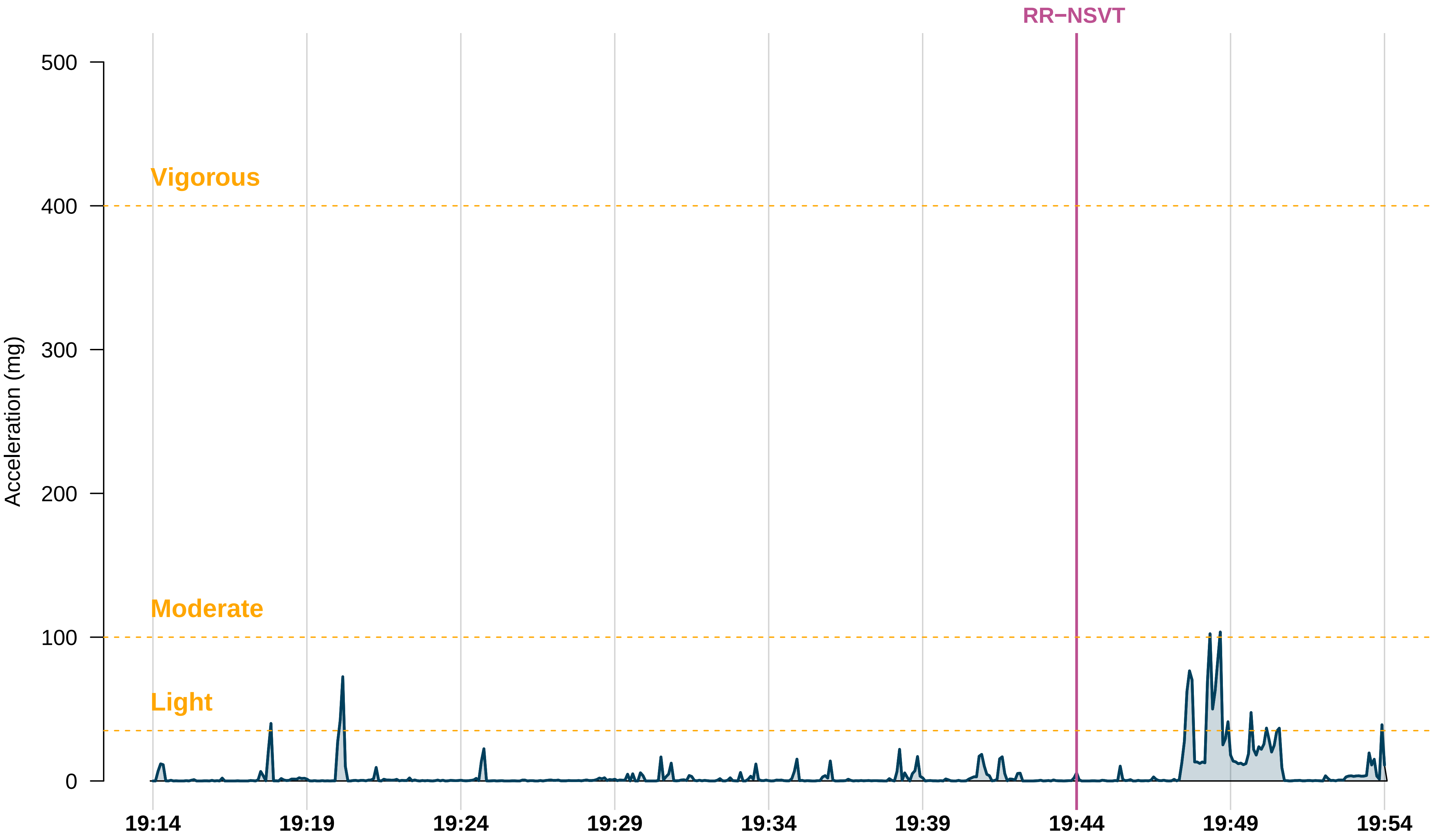

RR−NSVT
Vigorous
Moderate
Light
Acceleration (mg)
500
400
300
200
100
0
19:14
19:19
19:24
19:29
19:34
19:39
19:44
19:49
19:54

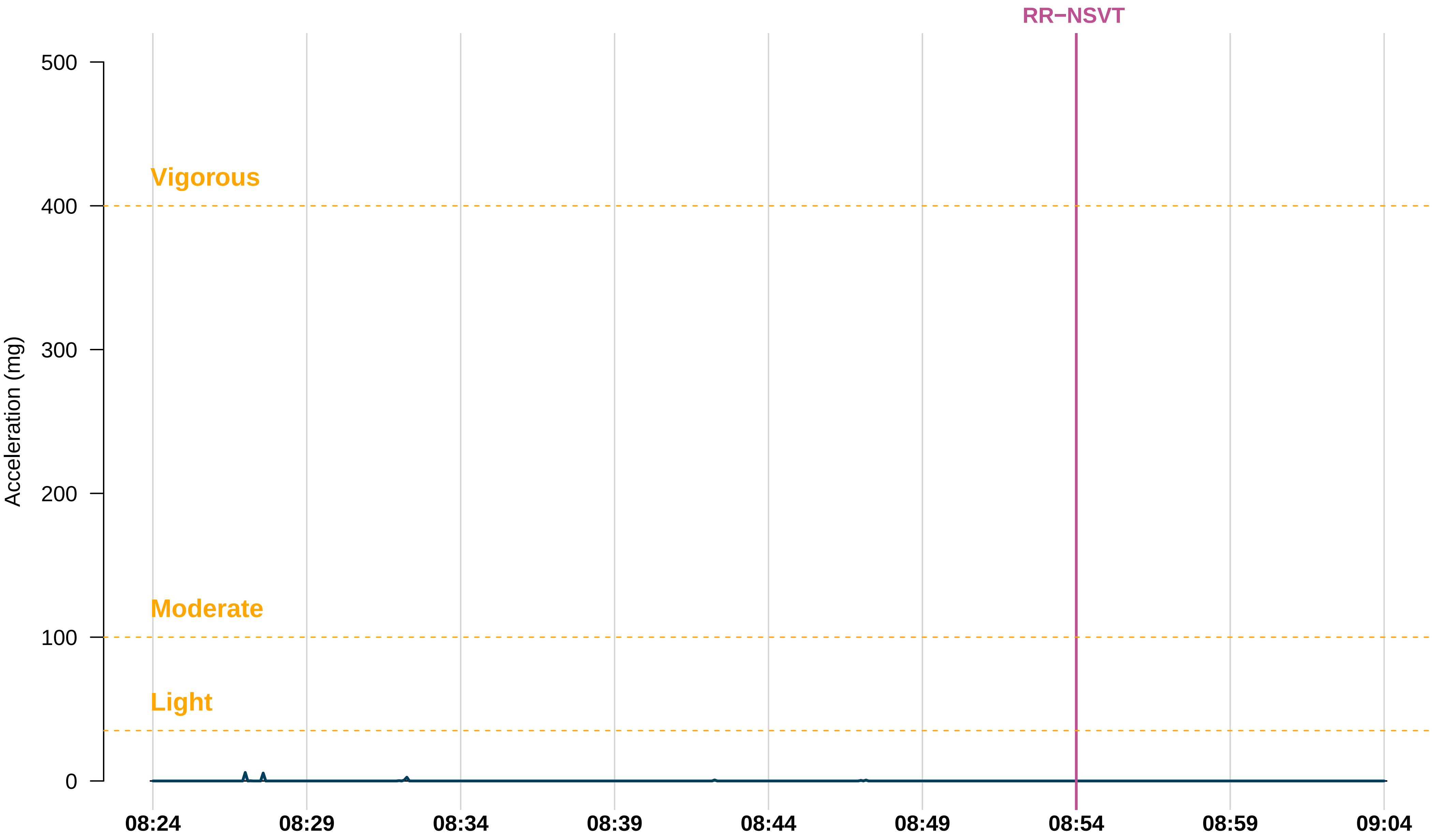
RR−NSVT
Vigorous
Moderate
Light
Acceleration (mg)
500
400
300
200
100
0
08:24
08:29
08:34
08:39
08:44
08:49
08:54
08:59
09:04

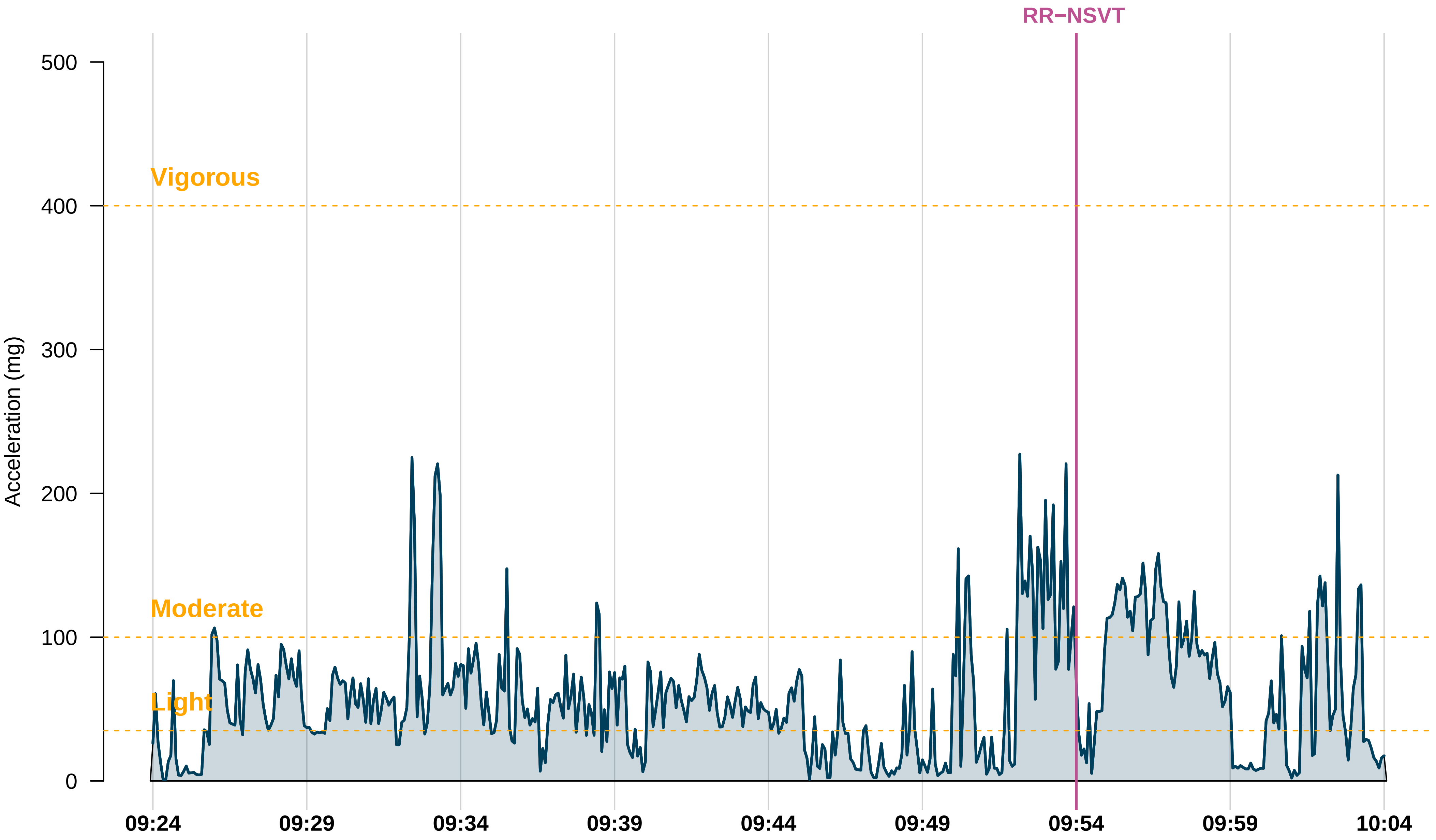
RR−NSVT
Vigorous
Moderate
Light
Acceleration (mg)
500
400
300
200
100
0
09:24
09:29
09:34
09:39
09:44
09:49
09:54
09:59
10:04

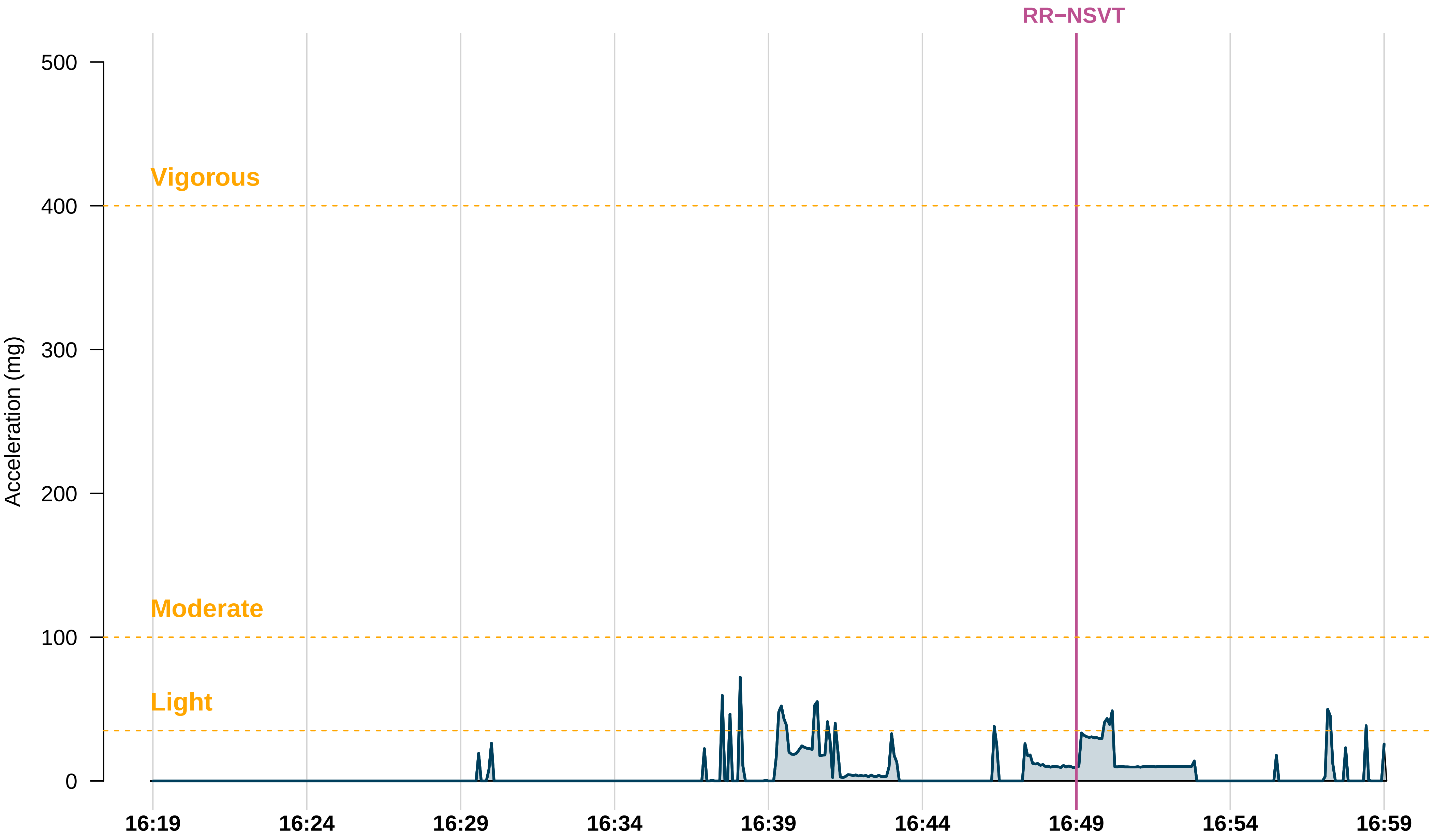
RR−NSVT
Vigorous
Moderate
Light
Acceleration (mg)
500
400
300
200
100
0
16:19
16:24
16:29
16:34
16:39
16:44
16:49
16:54
16:59

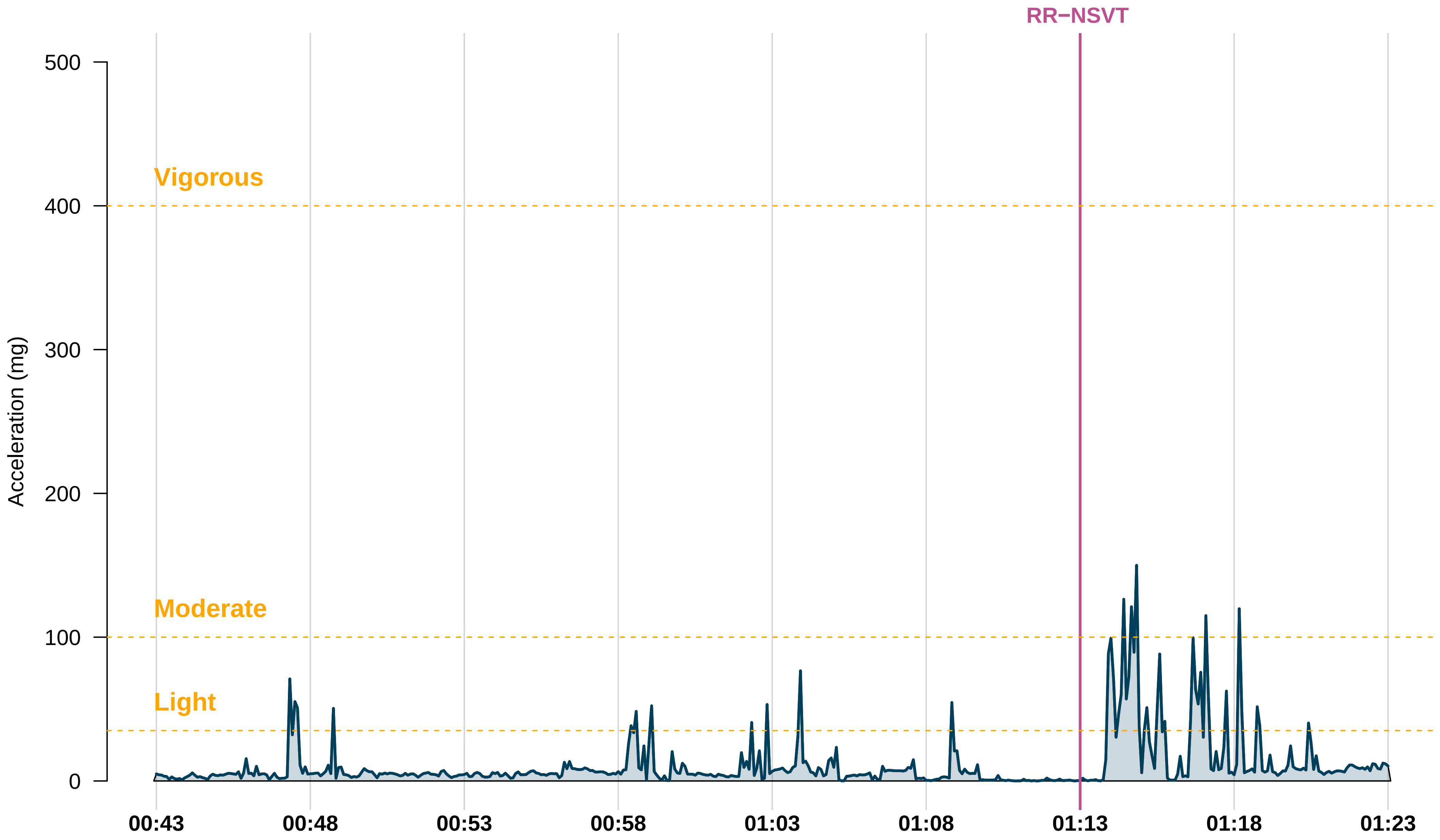

RR−NSVT
Vigorous
Moderate
Light
Acceleration (mg)
500
400
300
200
100
0
00:43
00:48
00:53
00:58
01:03
01:08
01:13
01:18
01:23

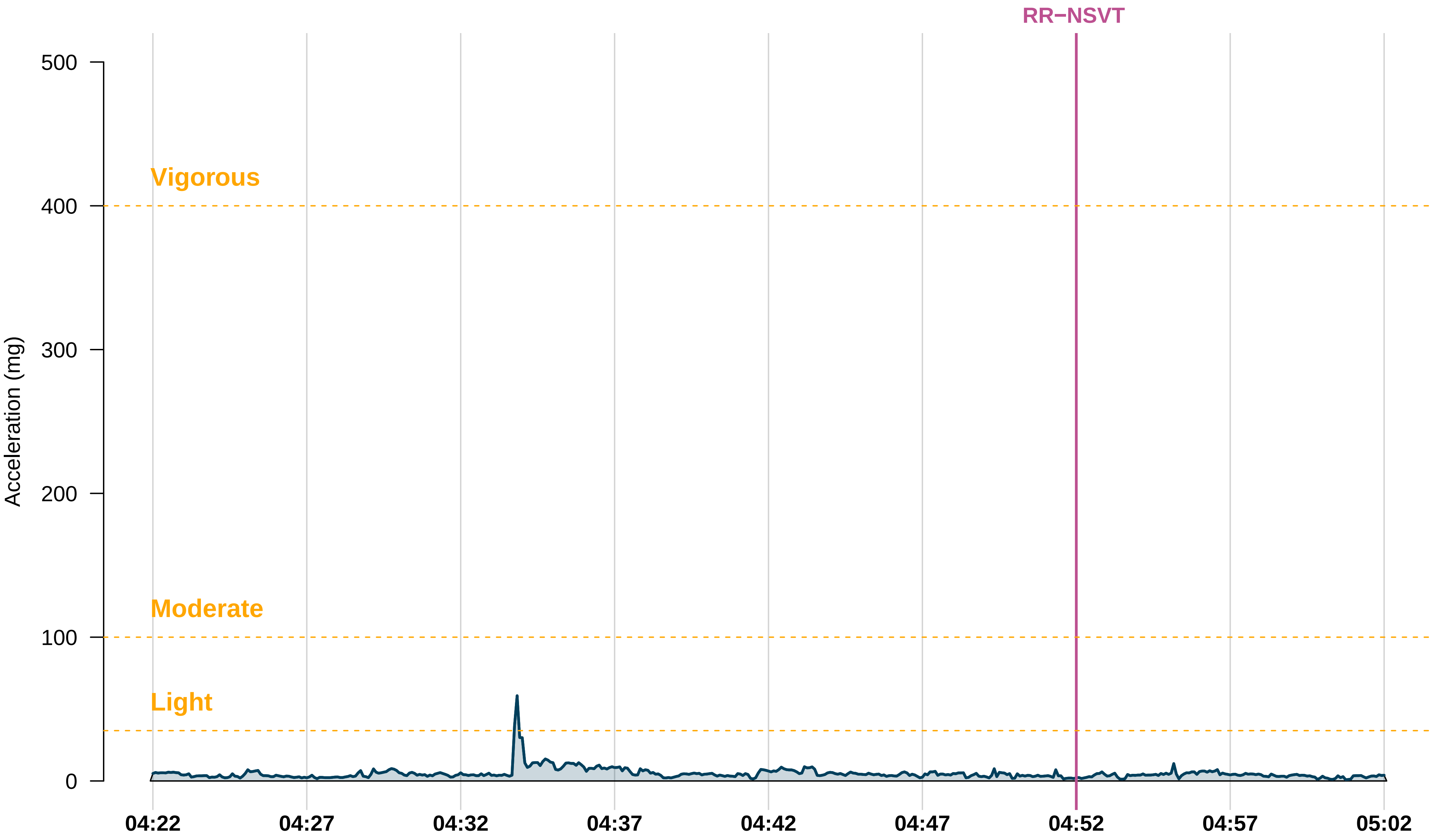

RR−NSVT
Vigorous
Moderate
Light
Acceleration (mg)
500
400
300
200
100
0
04:22
04:27
04:32
04:37
04:42
04:47
04:52
04:57
05:02

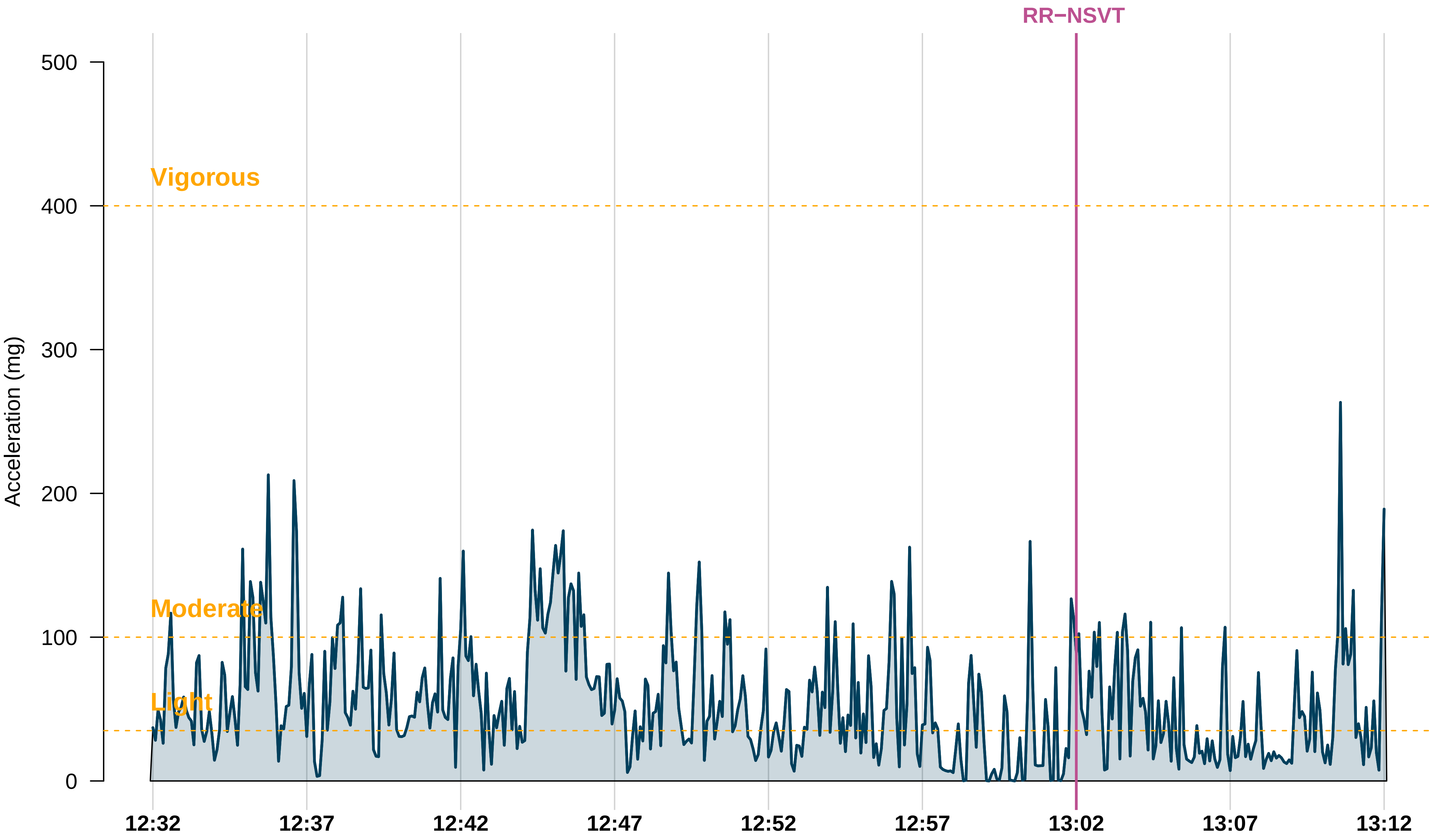

RR−NSVT
Vigorous
Moderate
Light
Acceleration (mg)
500
400
300
200
100
0
12:32
12:37
12:42
12:47
12:52
12:57
13:02
13:07
13:12

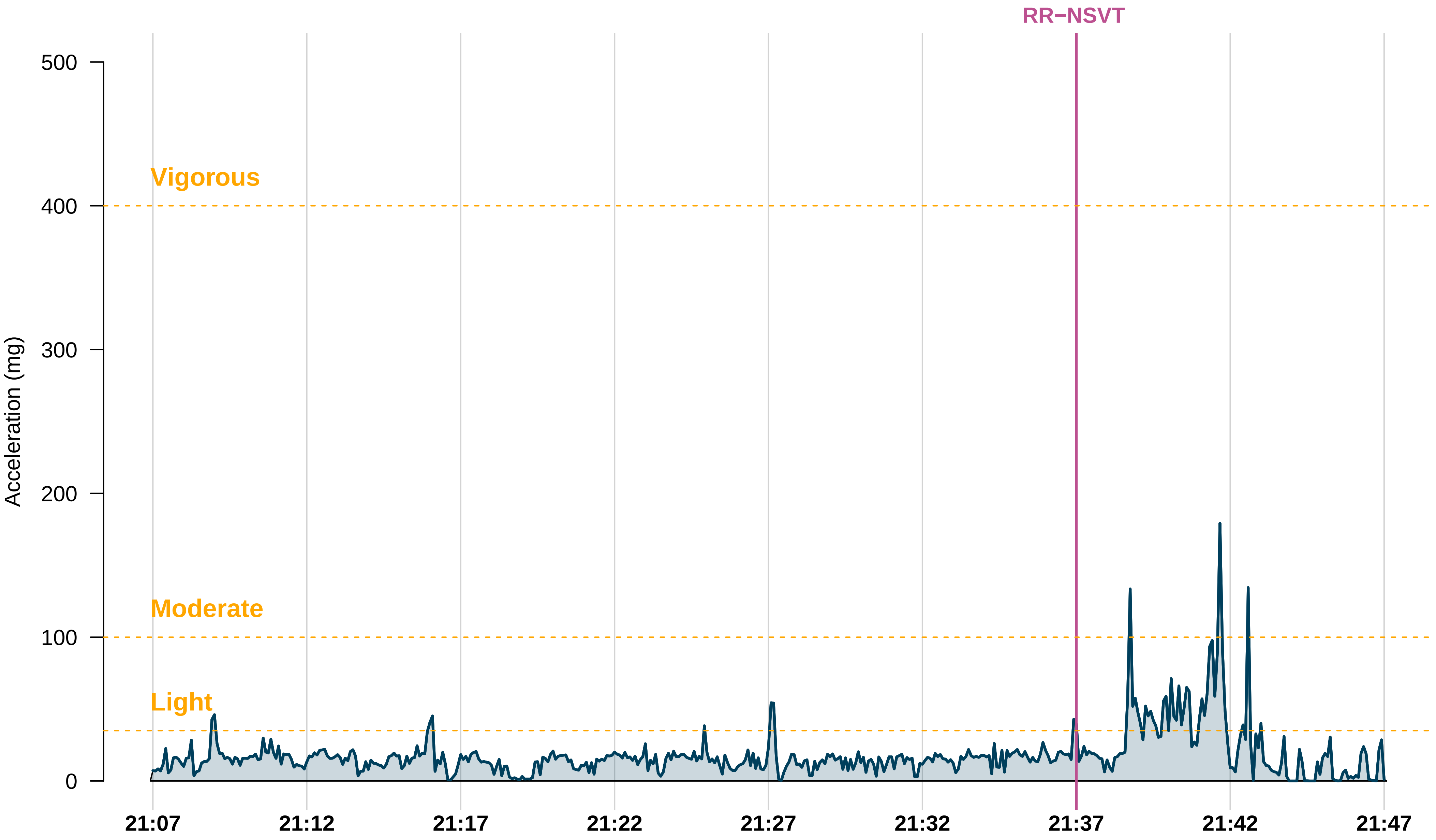
RR−NSVT
Vigorous
Moderate
Light
Acceleration (mg)
500
400
300
200
100
0
21:07
21:12
21:17
21:22
21:27
21:32
21:37
21:42
21:47

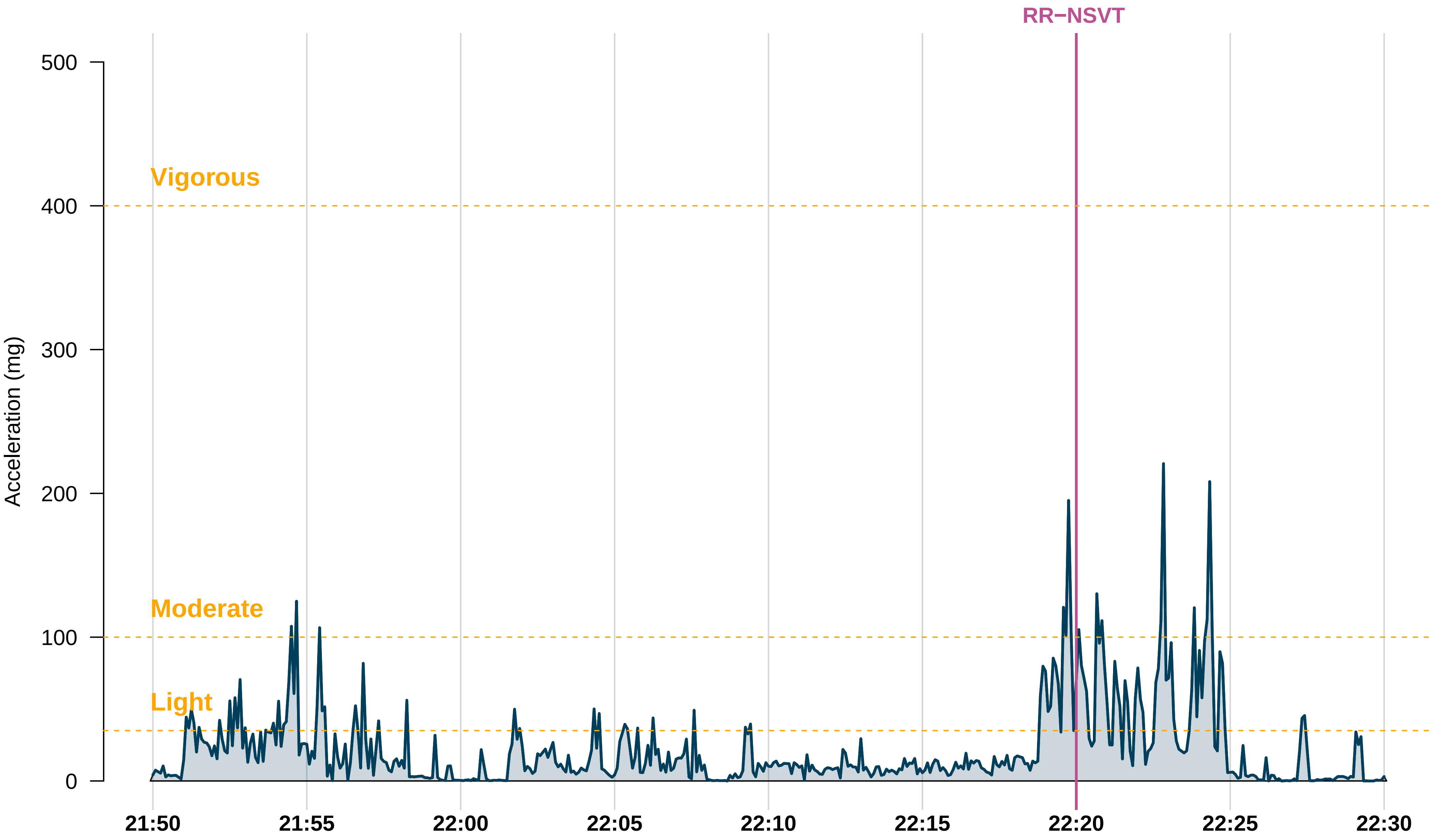
RR−NSVT
Vigorous
Moderate
Light
Acceleration (mg)
500
400
300
200
100
0
21:50
21:55
22:00
22:05
22:10
22:15
22:20
22:25
22:30

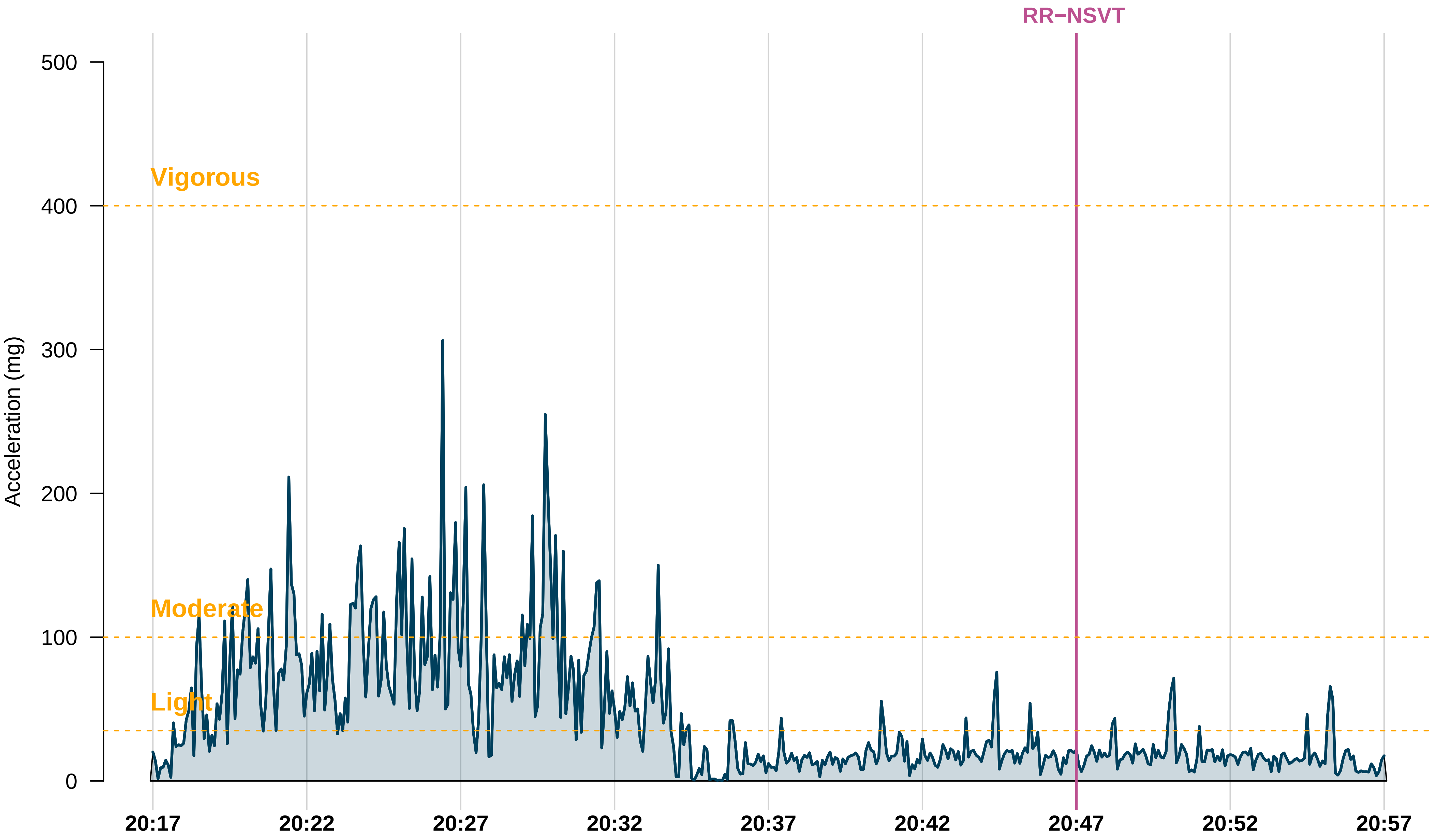

RR−NSVT
Vigorous
Moderate
Light
Acceleration (mg)
500
400
300
200
100
0
20:17
20:22
20:27
20:32
20:37
20:42
20:47
20:52
20:57

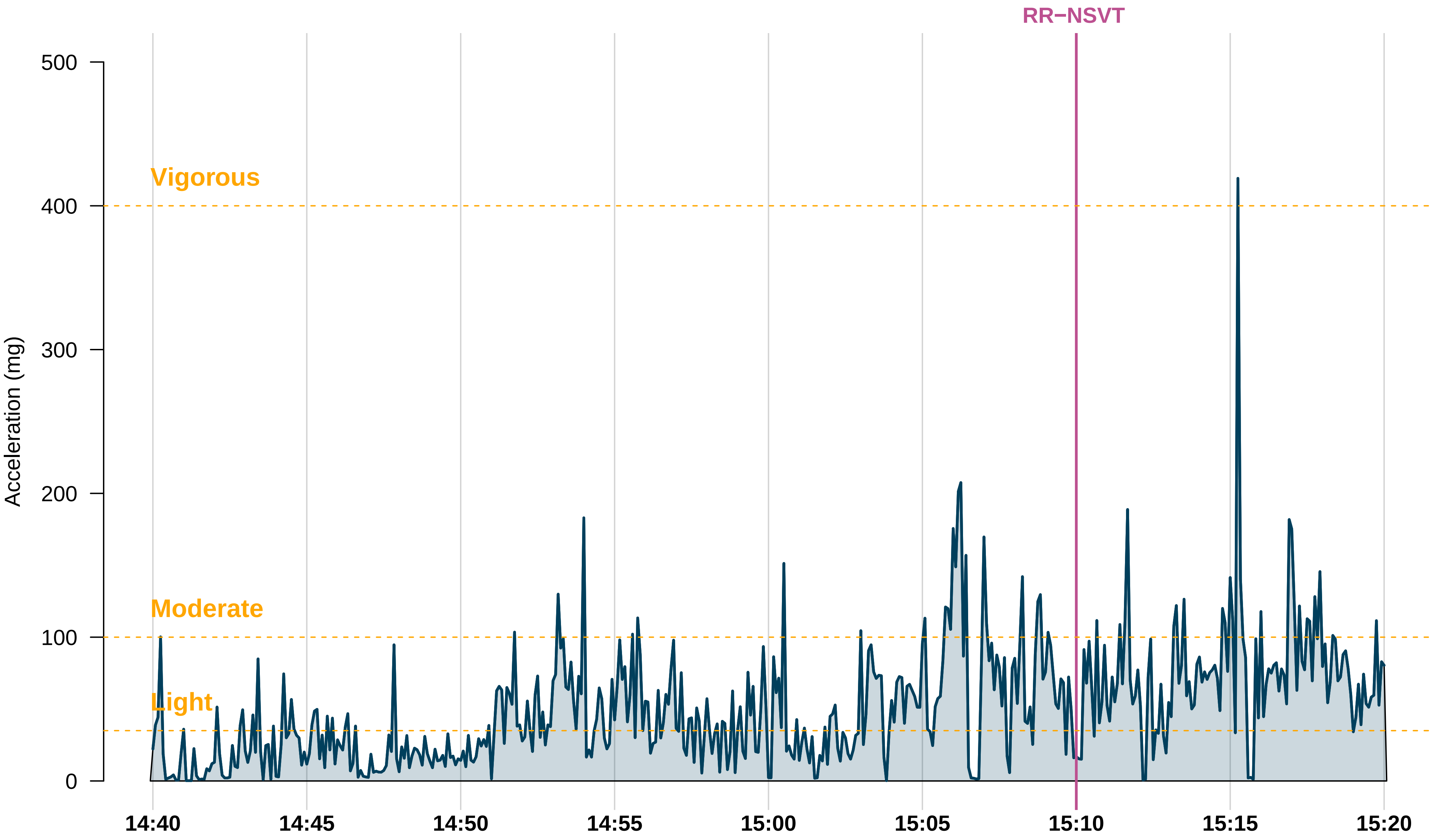

RR−NSVT
Vigorous
Moderate
Light
Acceleration (mg)
500
400
300
200
100
0
14:40
14:45
14:50
14:55
15:00
15:05
15:10
15:15
15:20

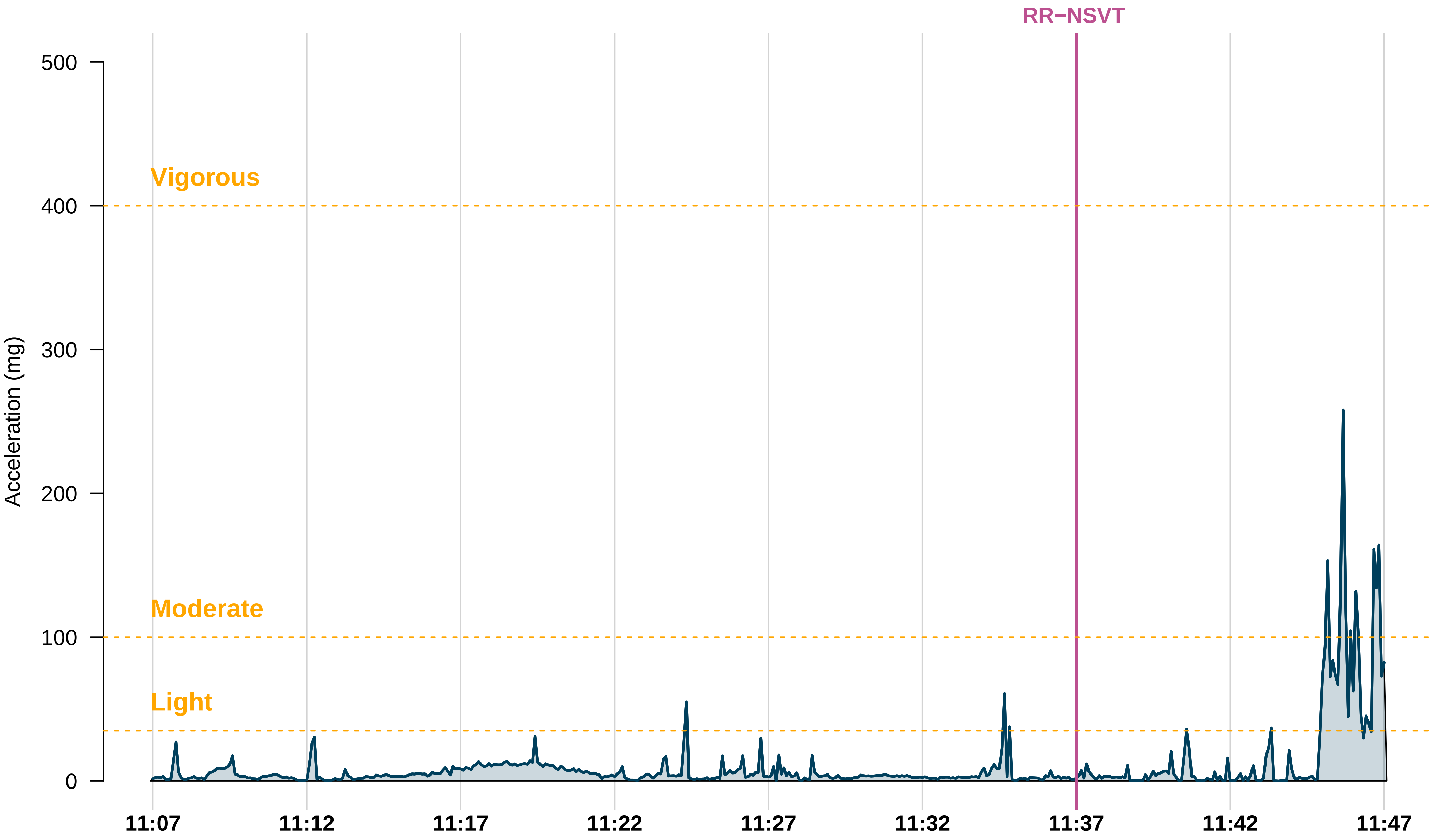
RR−NSVT
Vigorous
Moderate
Light
Acceleration (mg)
500
400
300
200
100
0
11:07
11:12
11:17
11:22
11:27
11:32
11:37
11:42
11:47

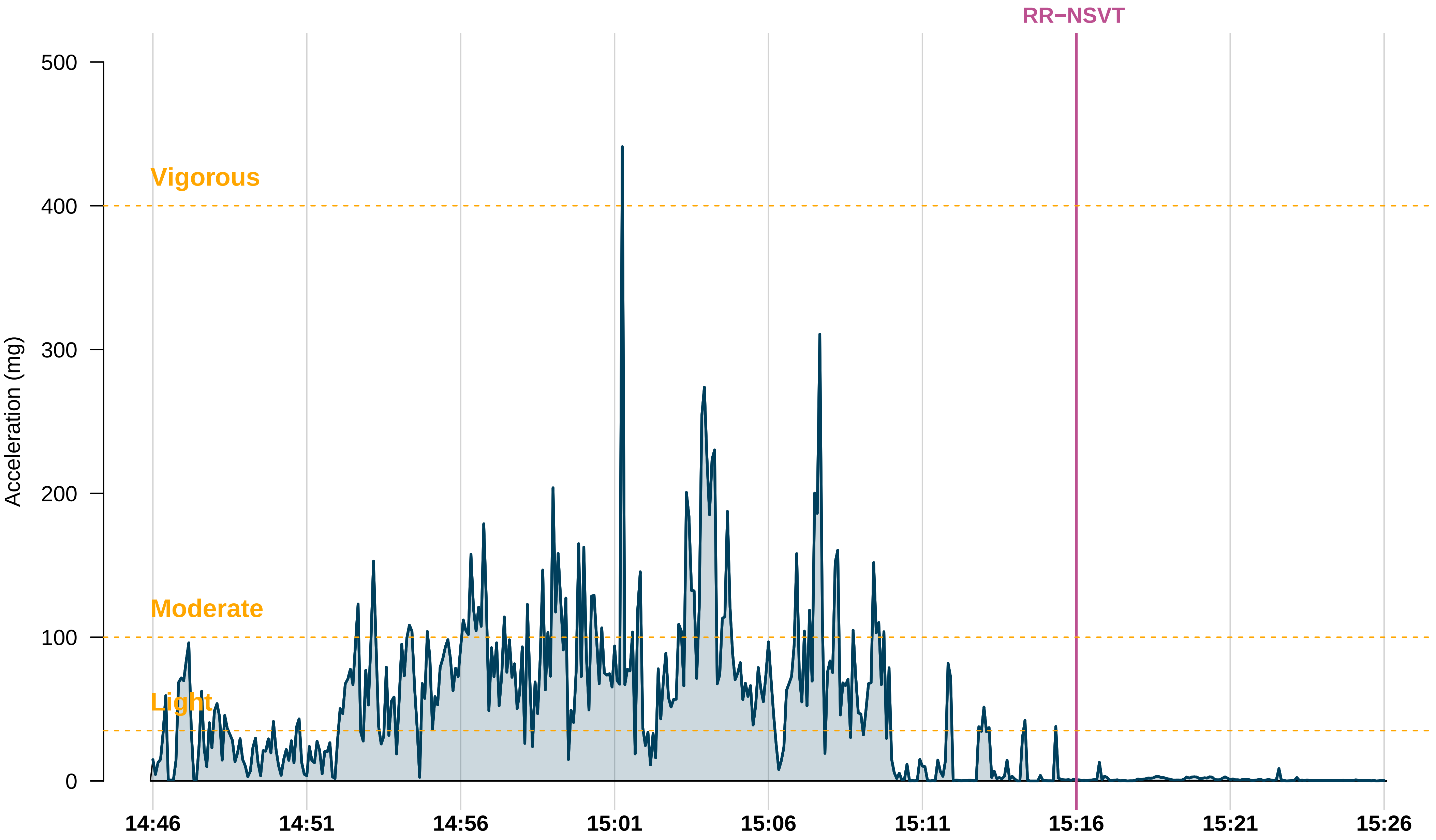

RR−NSVT
Vigorous
Moderate
Light
Acceleration (mg)
500
400
300
200
100
0
14:46
14:51
14:56
15:01
15:06
15:11
15:16
15:21
15:26

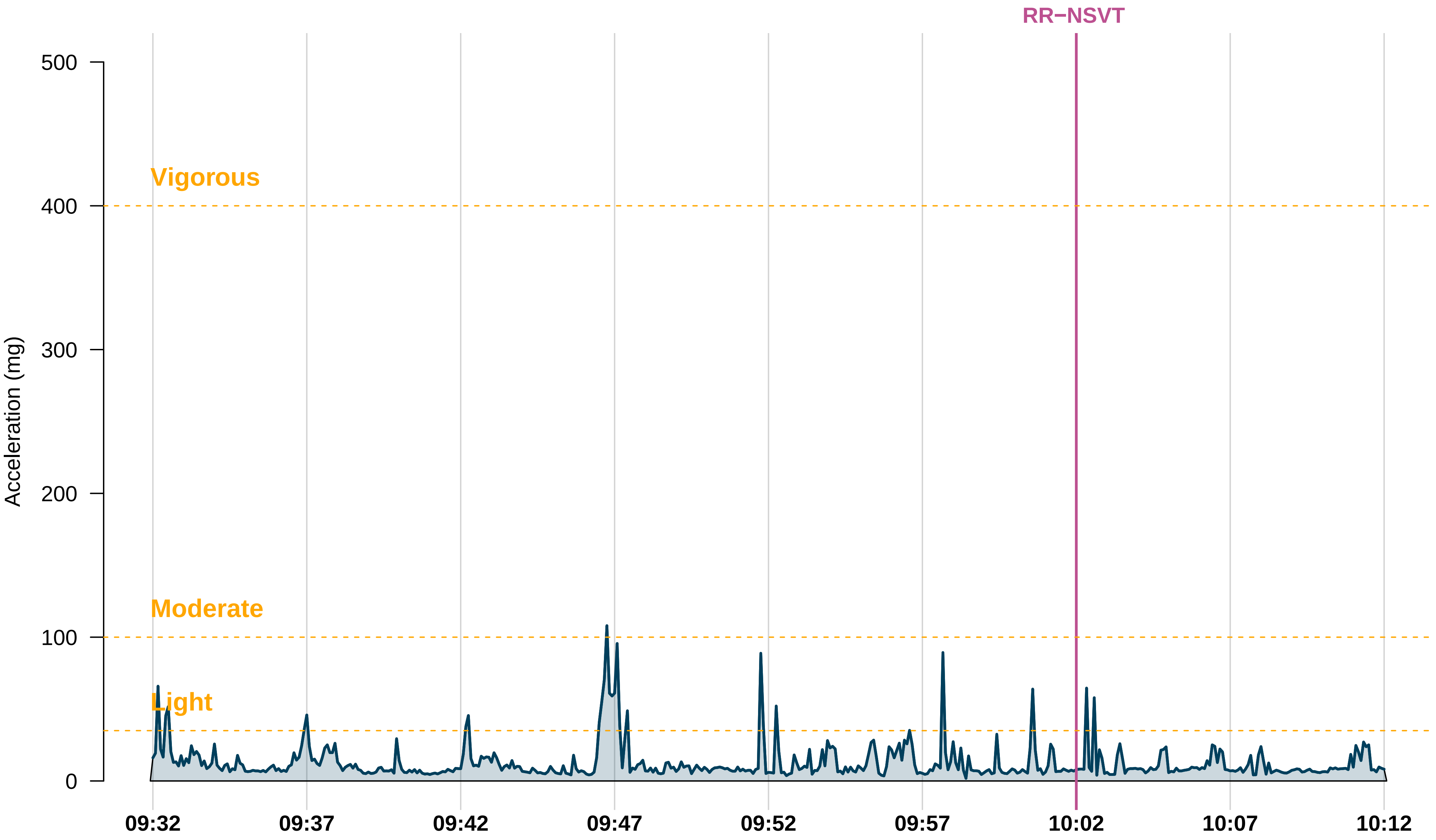

RR−NSVT
Vigorous
Moderate
Light
Acceleration (mg)
500
400
300
200
100
0
09:32
09:37
09:42
09:47
09:52
09:57
10:02
10:07
10:12

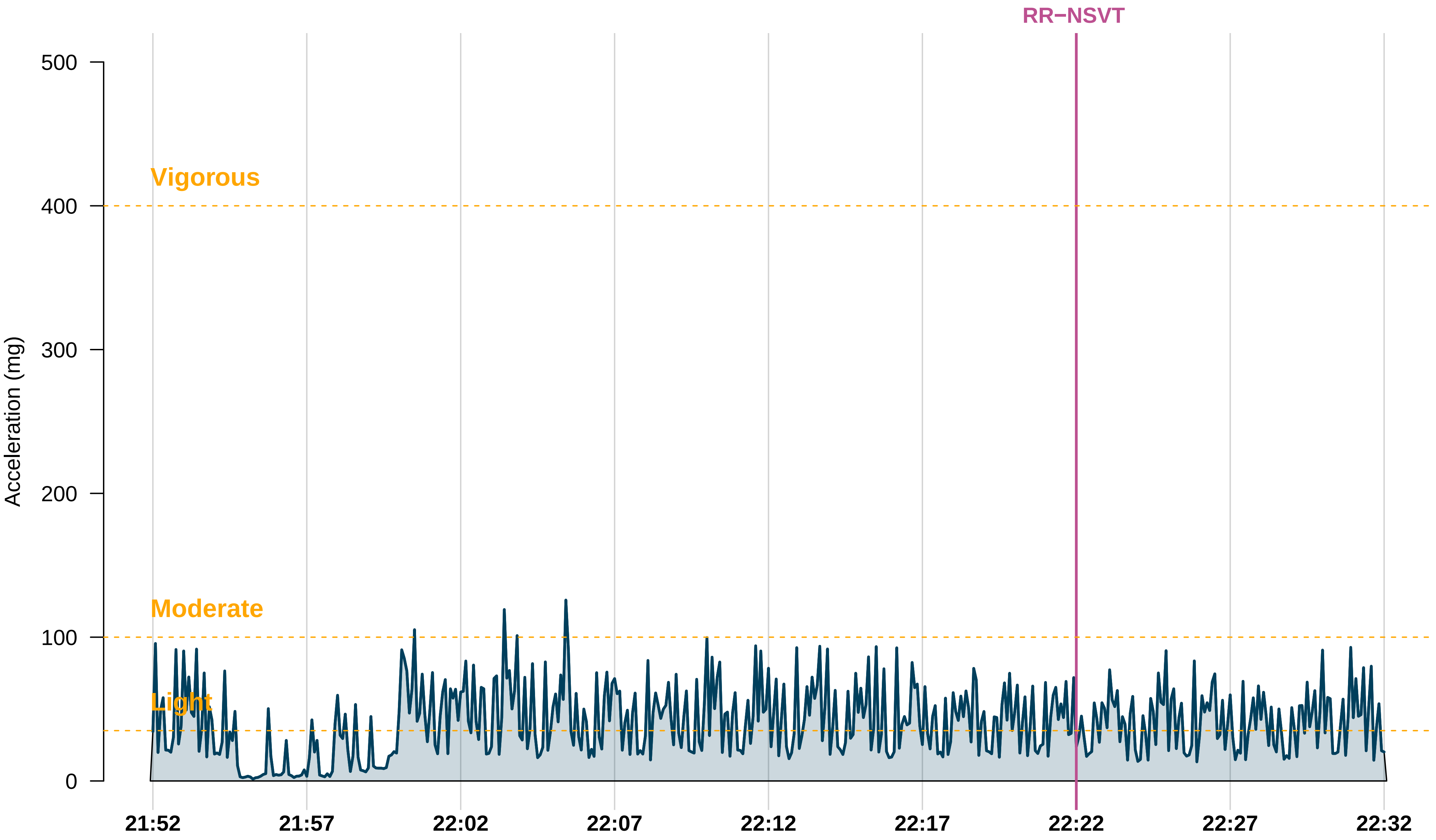
RR−NSVT
Vigorous
Moderate
Light
Acceleration (mg)
500
400
300
200
100
0
21:52
21:57
22:02
22:07
22:12
22:17
22:22
22:27
22:32

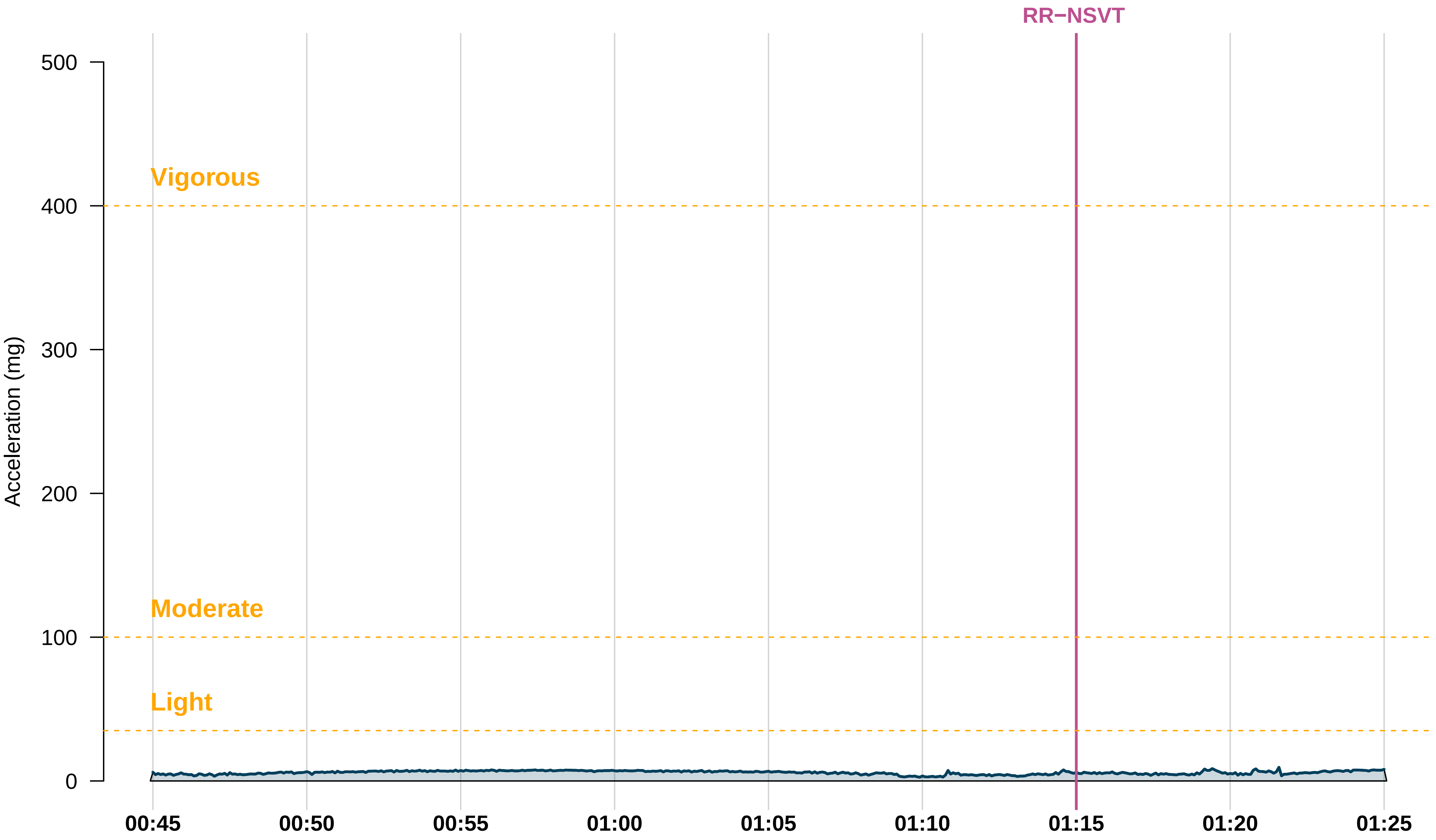
RR−NSVT
Vigorous
Moderate
Light
Acceleration (mg)
500
400
300
200
100
0
00:45
00:50
00:55
01:00
01:05
01:10
01:15
01:20
01:25

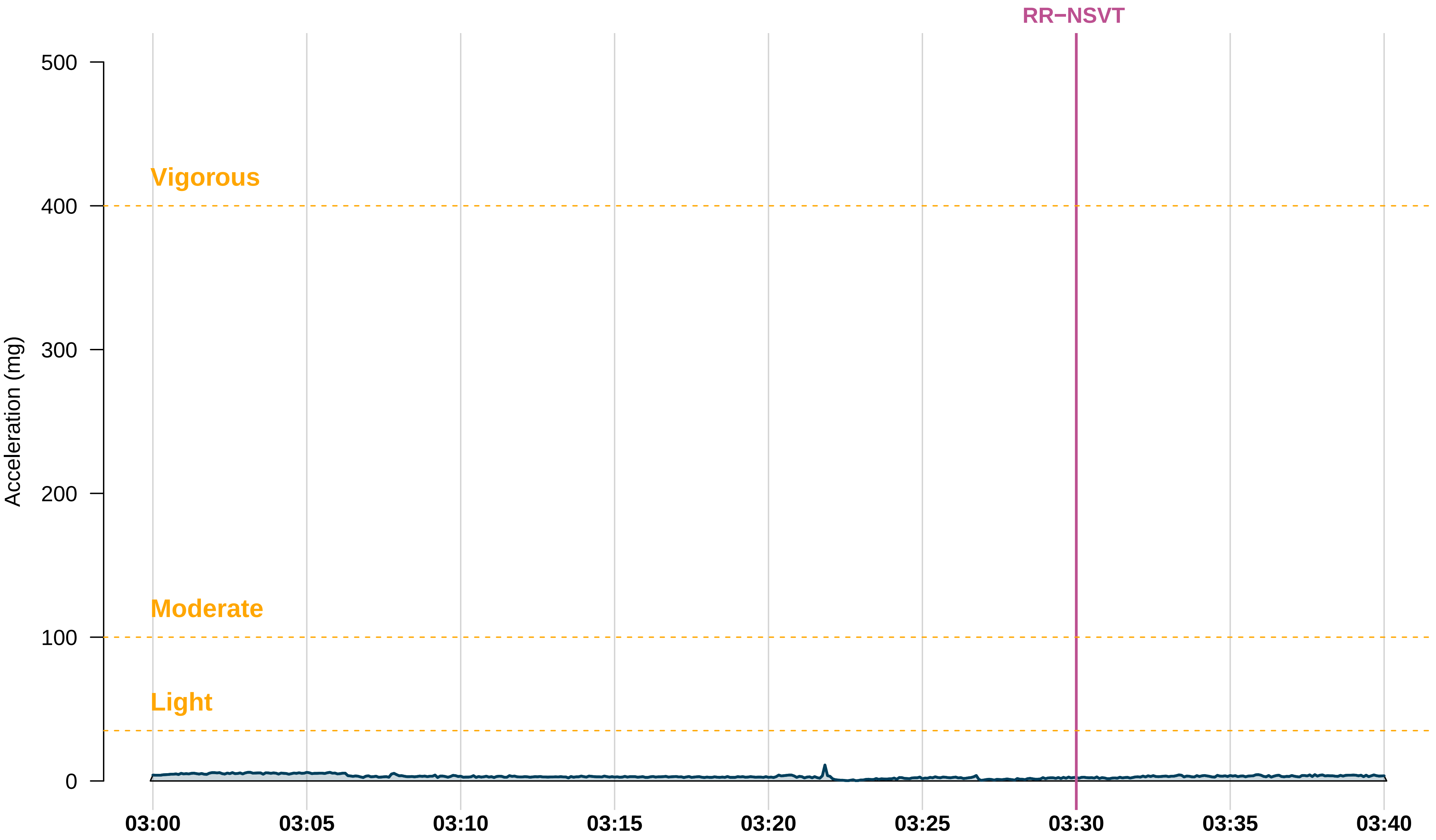
RR−NSVT
Acceleration (mg)
500
400
300
200
100
0
Vigorous
Moderate
Light
03:00
03:05
03:10
03:15
03:20
03:25
03:30
03:35
03:40

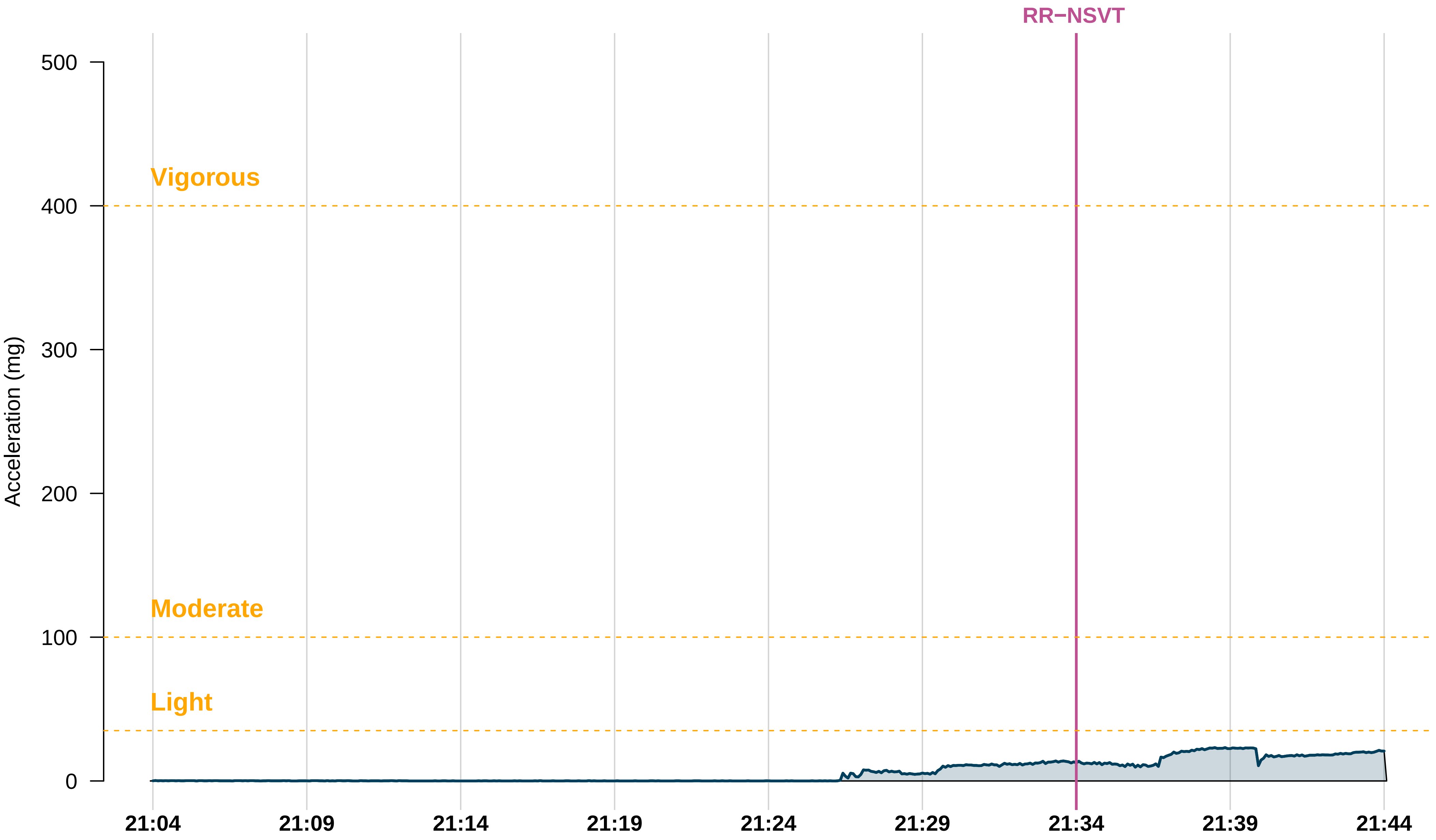

RR−NSVT
Vigorous
Moderate
Light
Acceleration (mg)
500
400
300
200
100
0
21:04
21:09
21:14
21:19
21:24
21:29
21:34
21:39
21:44

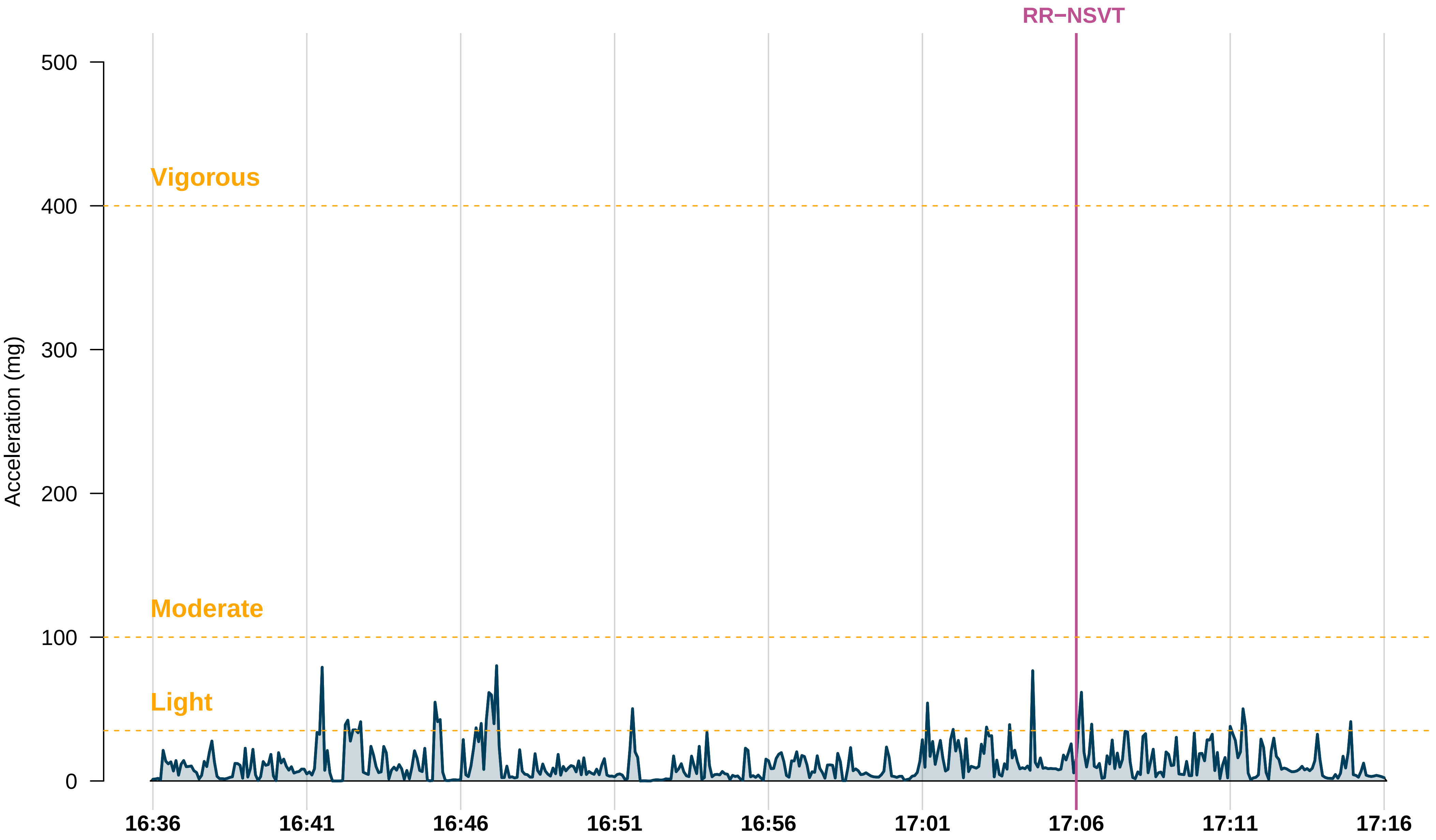
RR−NSVT
Vigorous
Moderate
Light
Acceleration (mg)
500
400
300
200
100
0
16:36
16:41
16:46
16:51
16:56
17:01
17:06
17:11
17:16

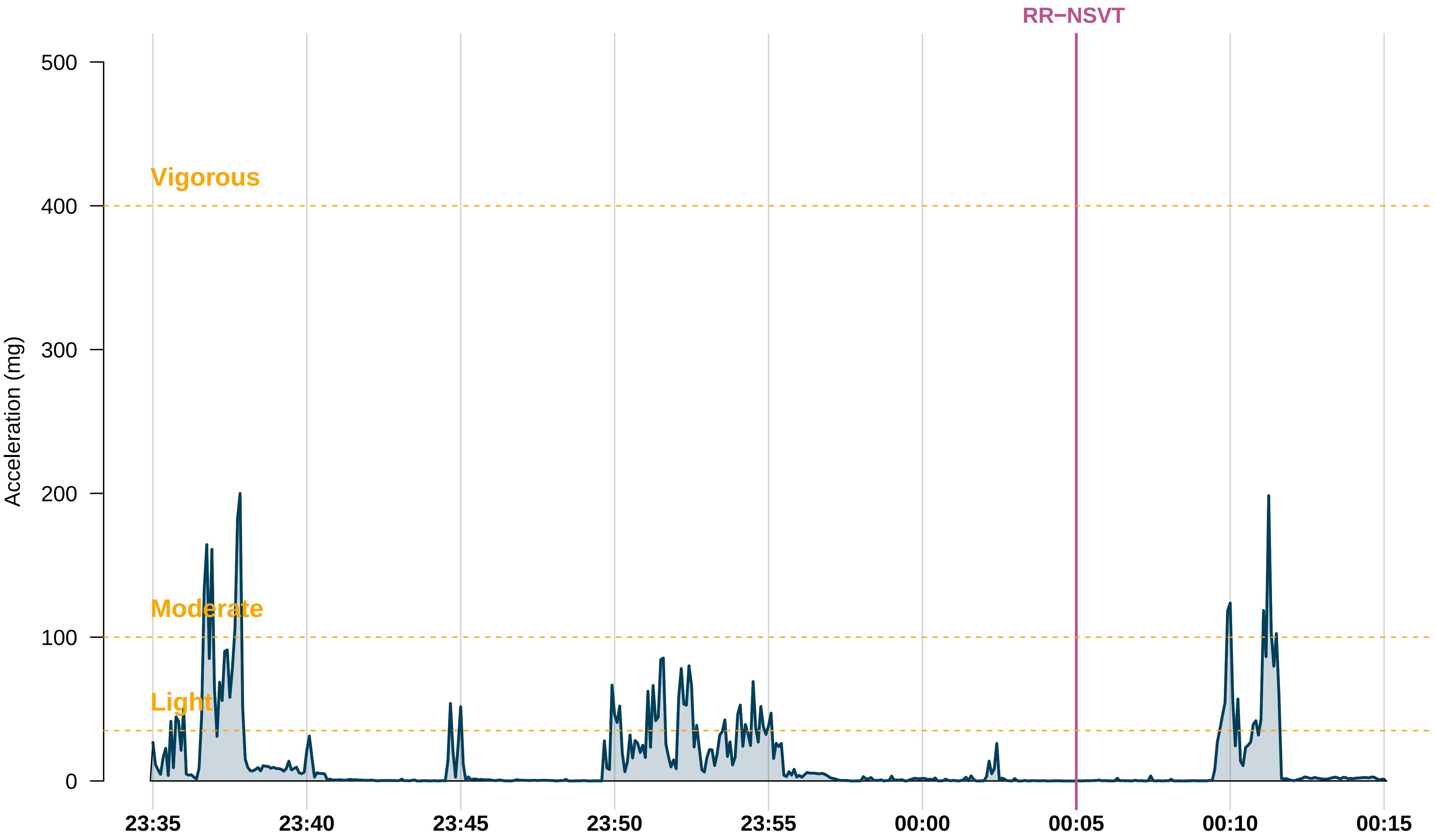
RR−NSVT
Vigorous
Moderate
Light
Acceleration (mg)
500
400
300
200
100
0
23:35
23:40
23:45
23:50
23:55
00:00
00:05
00:10
00:15

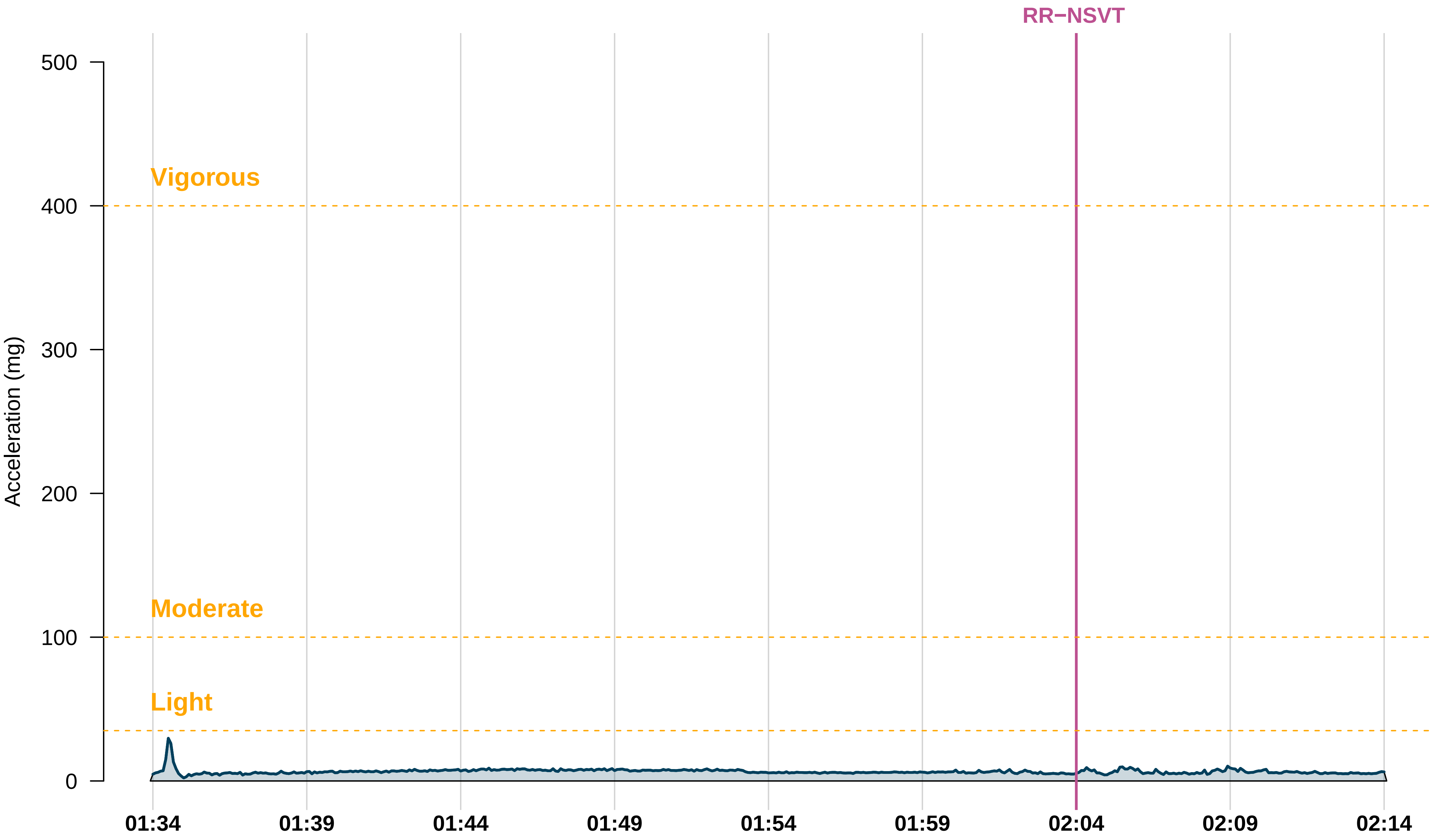

RR−NSVT
Vigorous
Moderate
Light
Acceleration (mg)
500
400
300
200
100
0
01:34
01:39
01:44
01:49
01:54
01:59
02:04
02:09
02:14

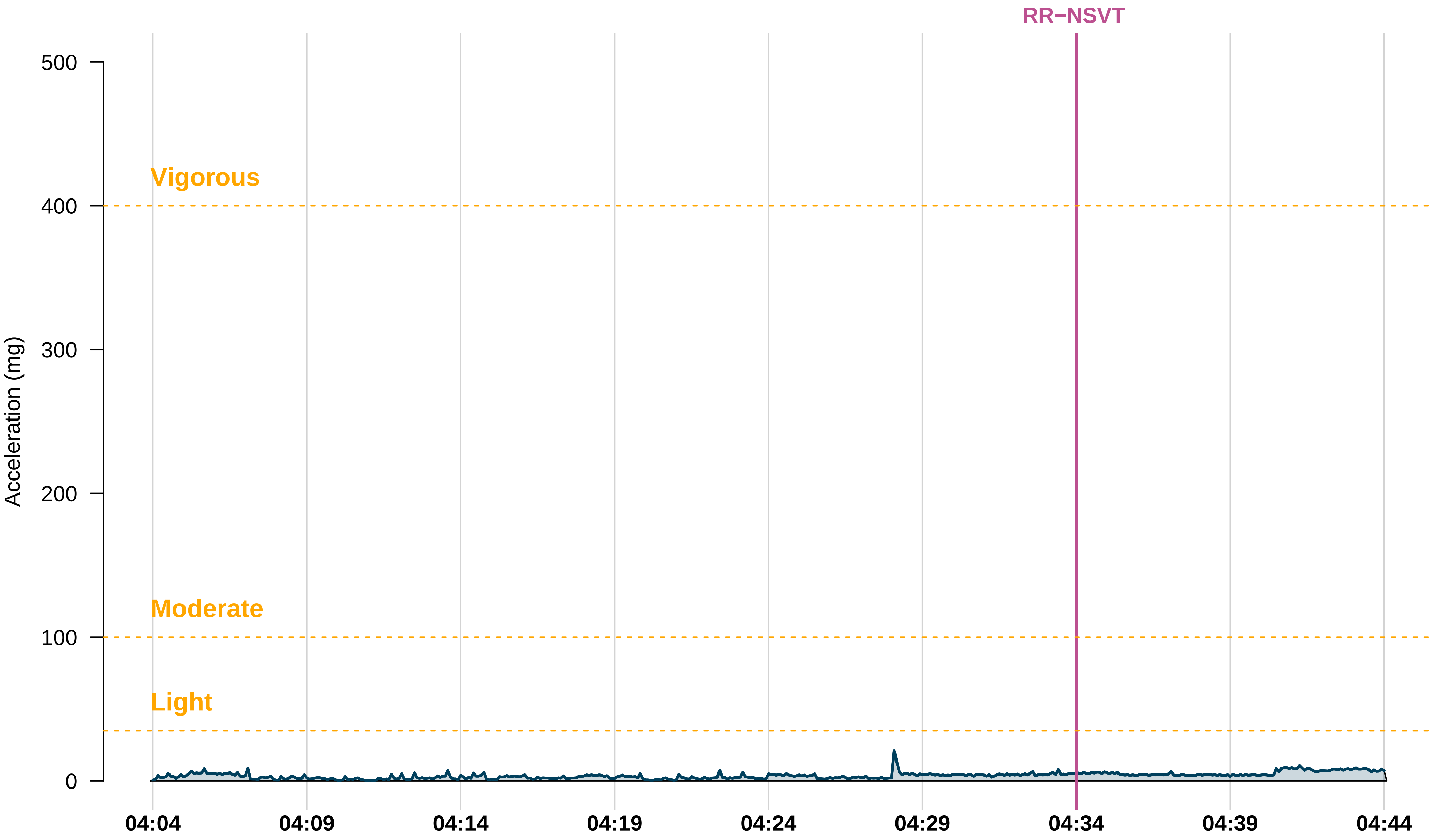

RR−NSVT
Vigorous
Moderate
Light
Acceleration (mg)
500
400
300
200
100
0
04:04
04:09
04:14
04:19
04:24
04:29
04:34
04:39
04:44

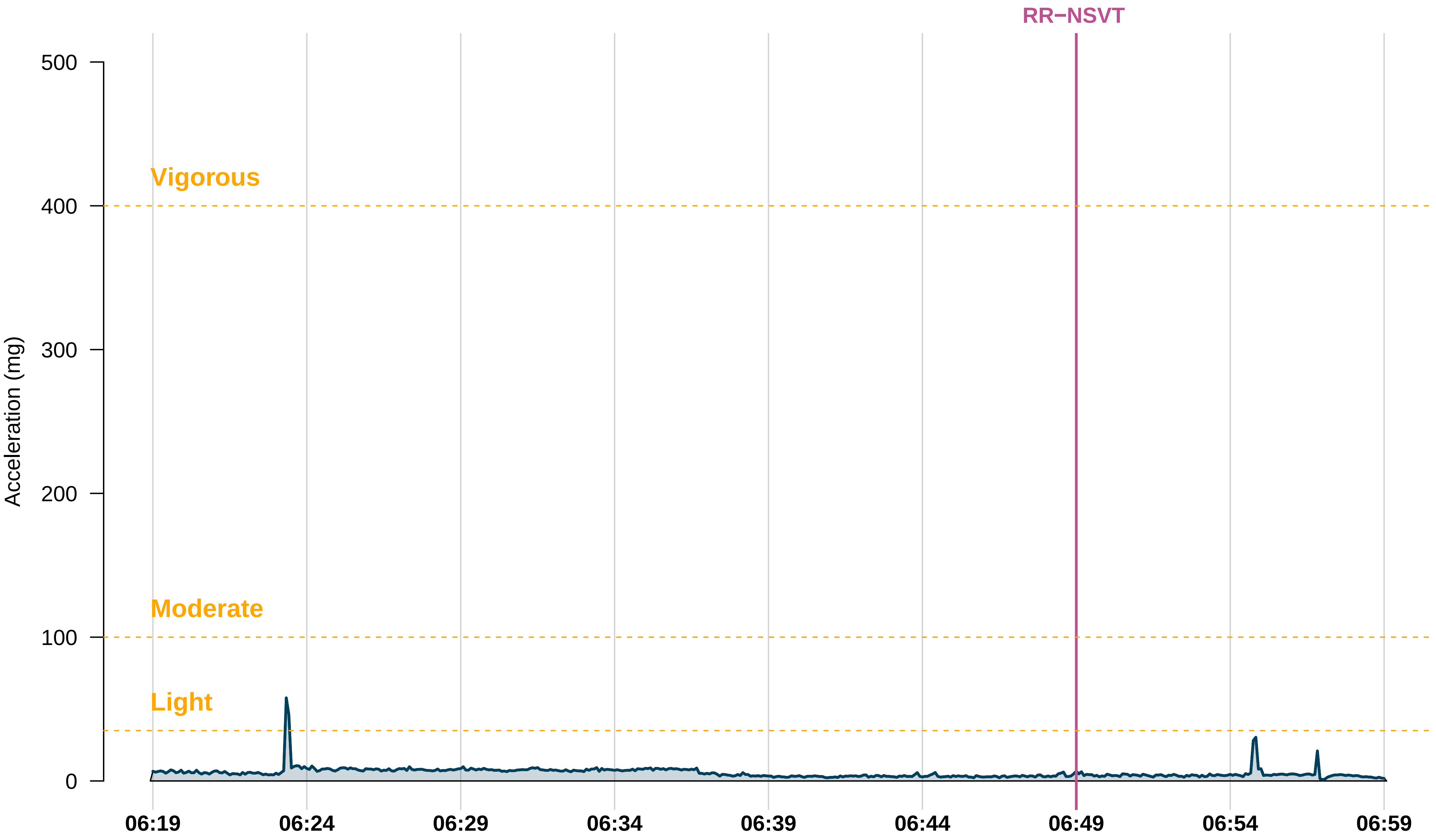

RR−NSVT
Vigorous
Moderate
Light
Acceleration (mg)
500
400
300
200
100
0
06:19
06:24
06:29
06:34
06:39
06:44
06:49
06:54
06:59

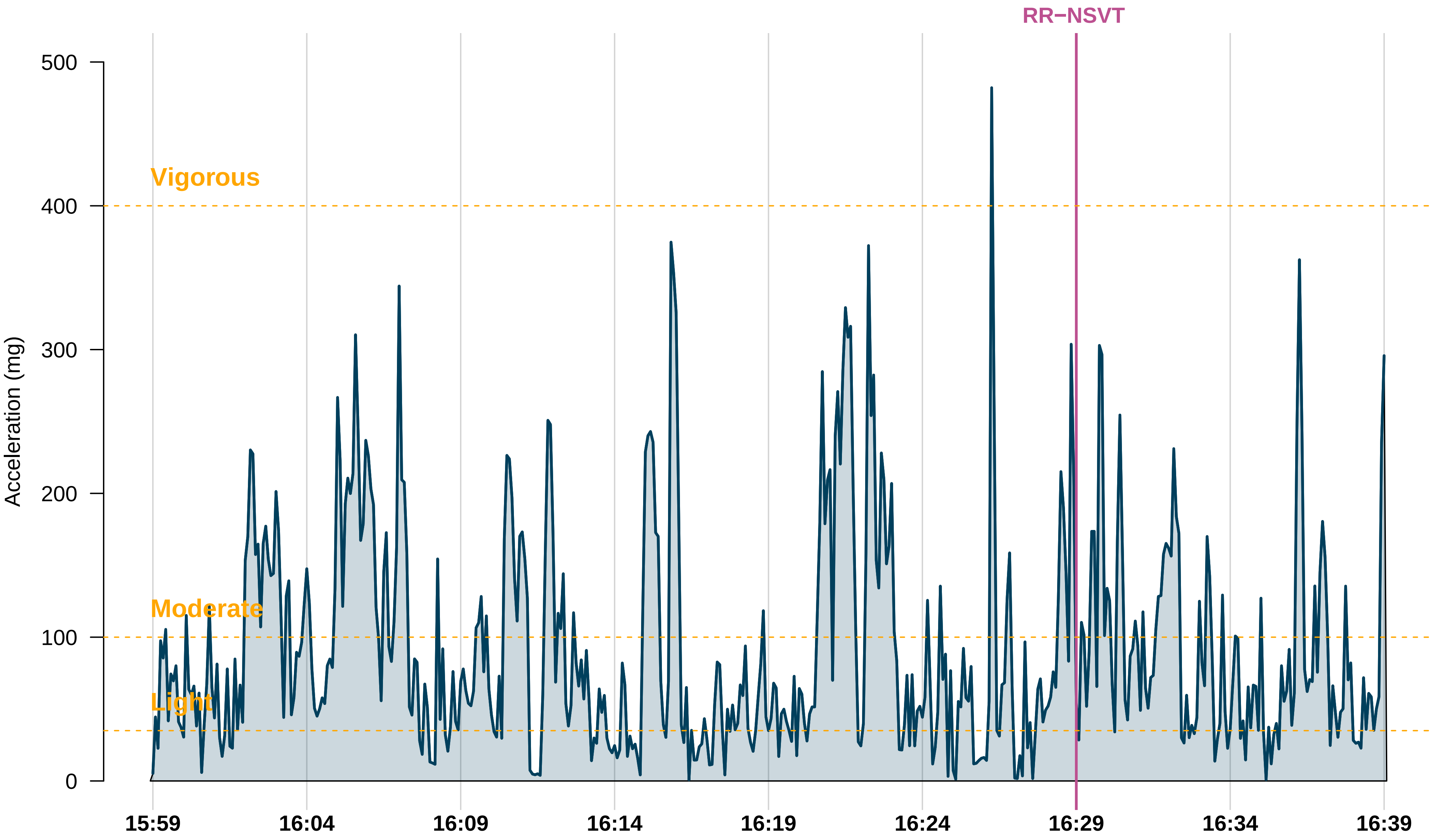

RR−NSVT
Vigorous
Moderate
Light
Acceleration (mg)
500
400
300
200
100
0
15:59
16:04
16:09
16:14
16:19
16:24
16:29
16:34
16:39

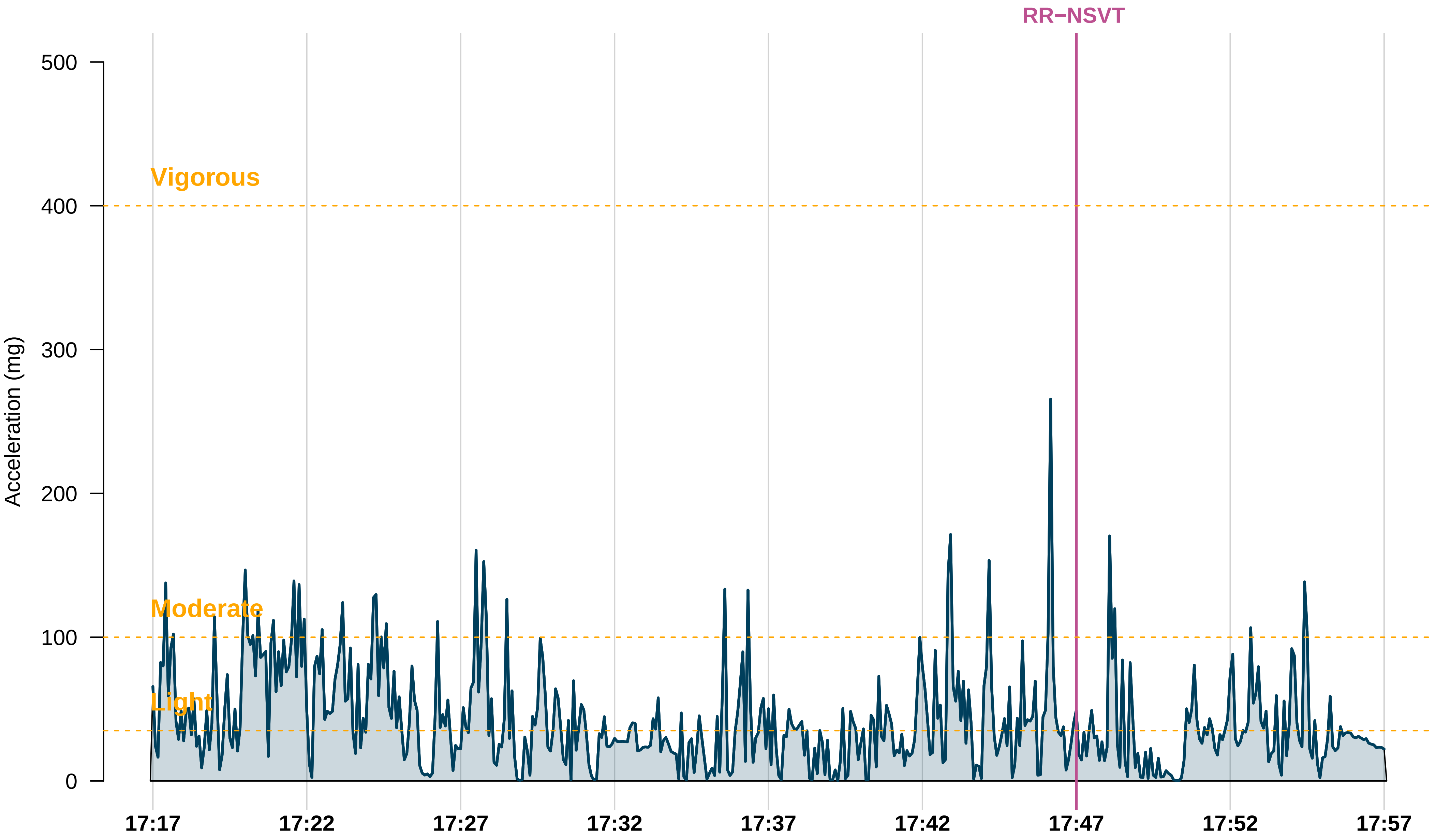

RR−NSVT
Vigorous
Moderate
Light
Acceleration (mg)
500
400
300
200
100
0
17:17
17:22
17:27
17:32
17:37
17:42
17:47
17:52
17:57

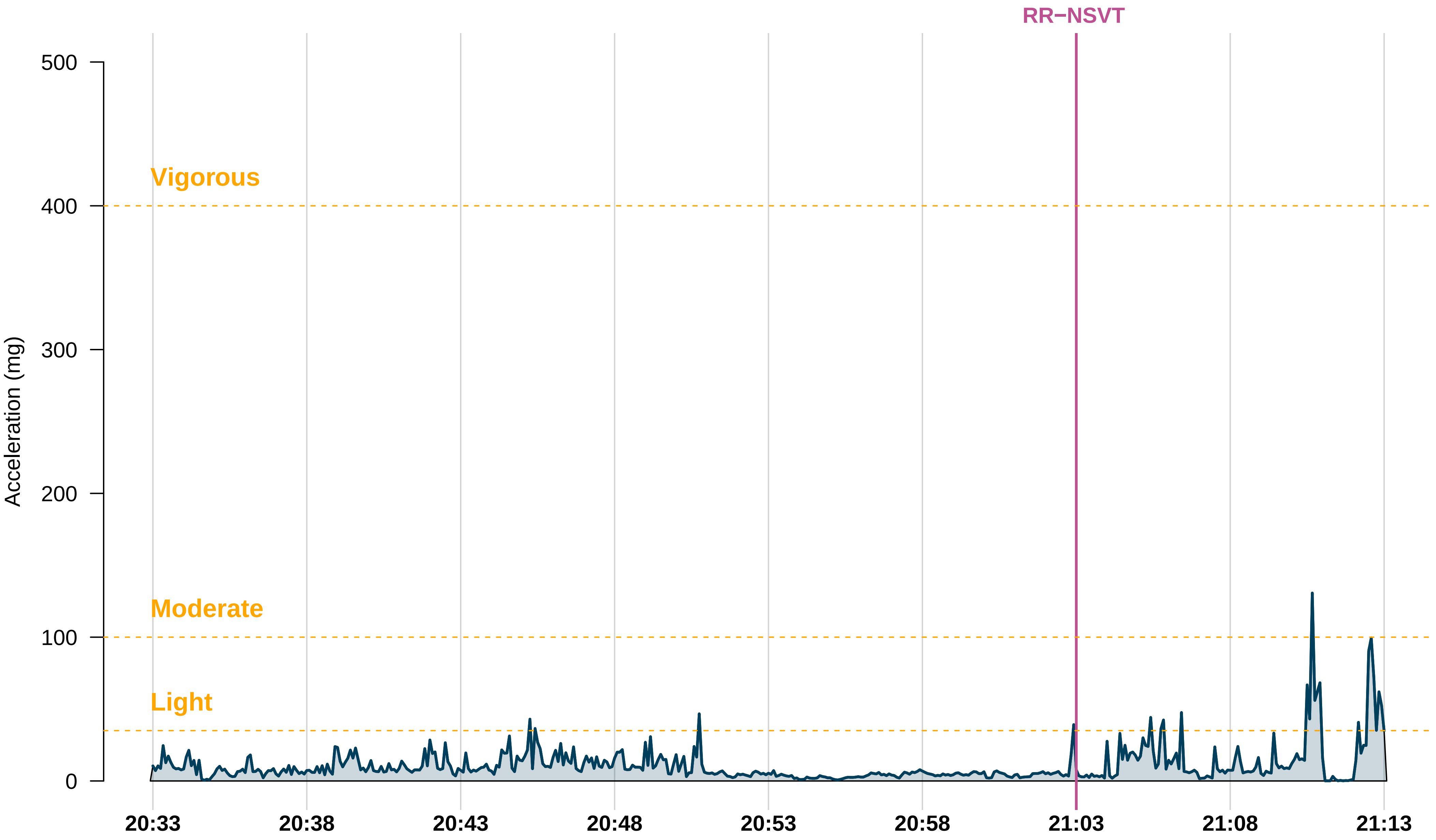

RR−NSVT
Vigorous
Moderate
Light
Acceleration (mg)
500
400
300
200
100
0
20:33
20:38
20:43
20:48
20:53
20:58
21:03
21:08
21:13

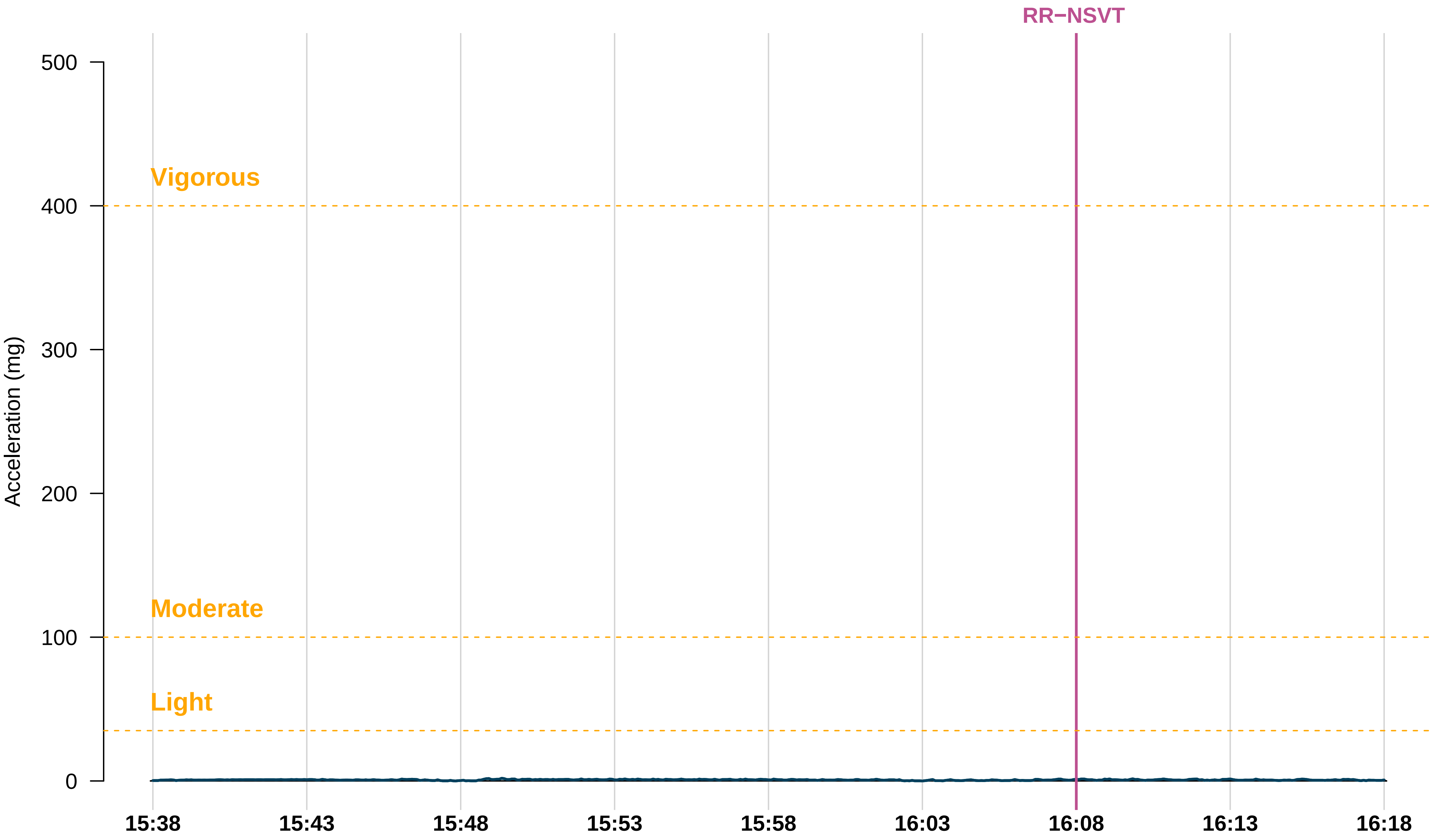
RR−NSVT
Acceleration (mg)
500
400
300
200
100
0
Vigorous
Moderate
Light
15:38
15:43
15:48
15:53
15:58
16:03
16:08
16:13
16:18

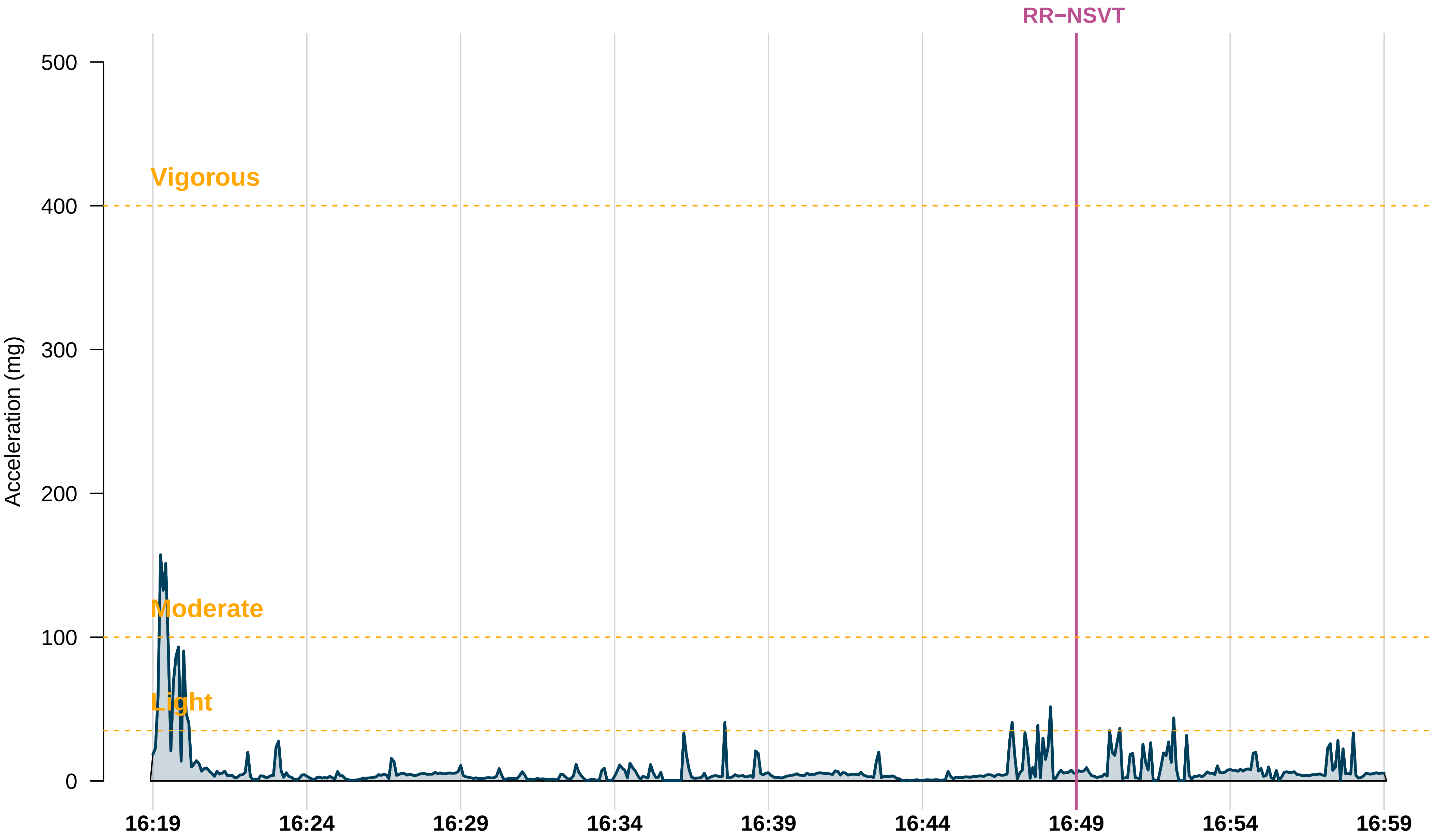

RR−NSVT
Vigorous
Moderate
Light
Acceleration (mg)
500
400
300
200
100
0
16:19
16:24
16:29
16:34
16:39
16:44
16:49
16:54
16:59

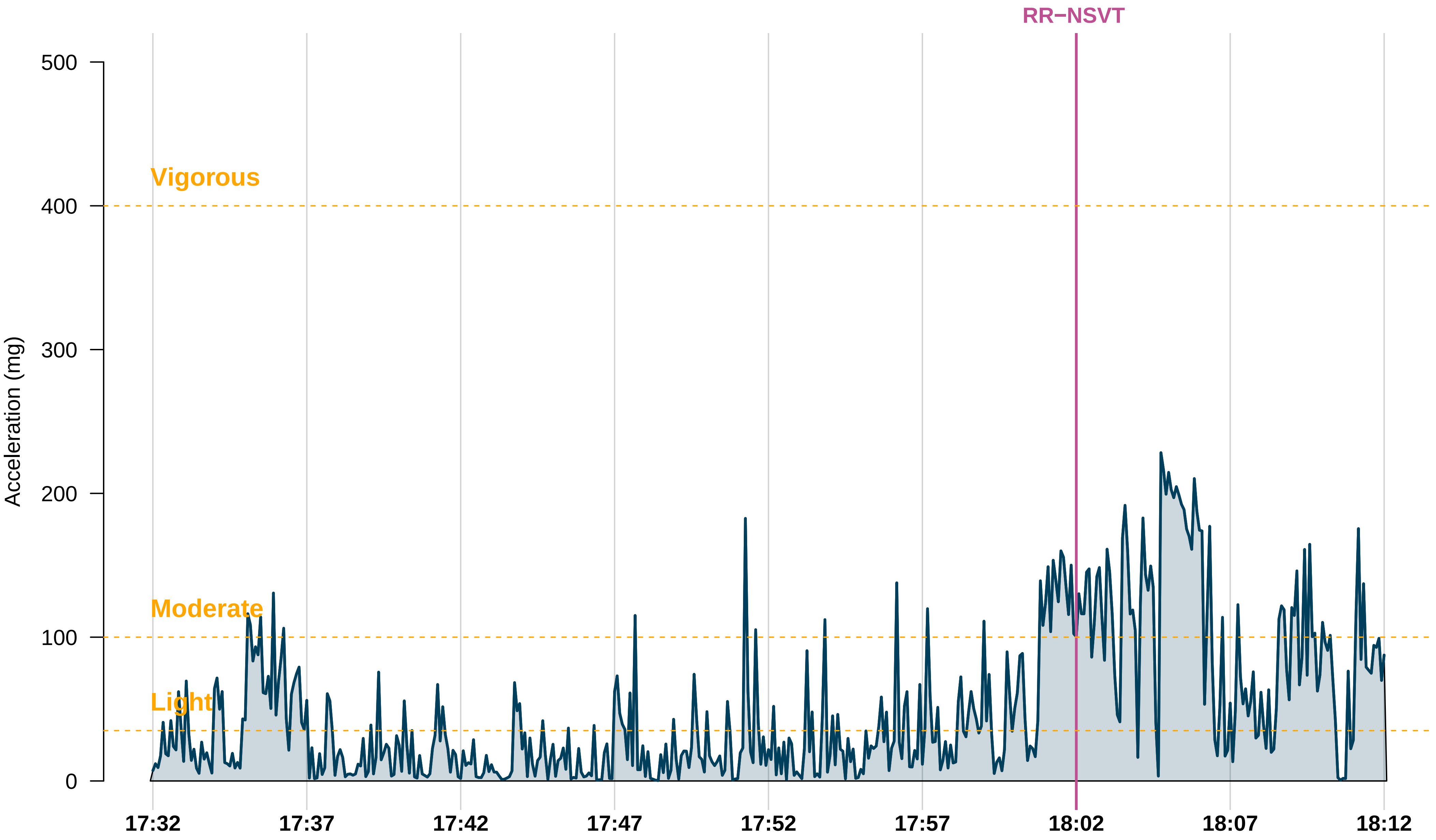

RR−NSVT
Vigorous
Moderate
Light
Acceleration (mg)
0
100
200
300
400
500
17:32
17:37
17:42
17:47
17:52
17:57
18:02
18:07
18:12

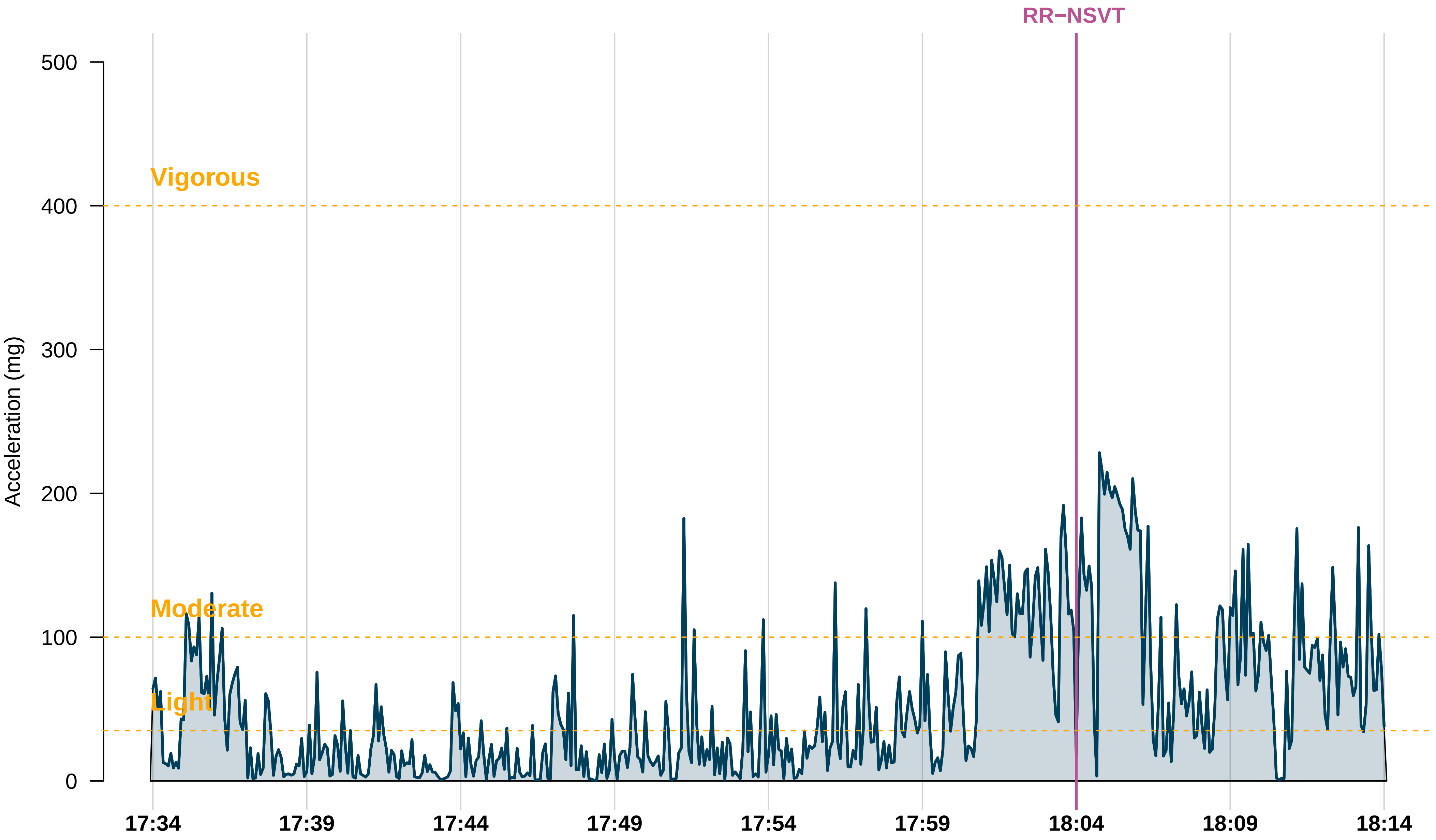

RR−NSVT
Vigorous
Moderate
Light
Acceleration (mg)
500
400
300
200
100
0
17:34
17:39
17:44
17:49
17:54
17:59
18:04
18:09
18:14
